# Altered cellular and humoral immune responses following SARS-CoV-2 mRNA vaccination in patients with multiple sclerosis on anti-CD20 therapy

**DOI:** 10.1101/2021.06.23.21259389

**Authors:** Sokratis A. Apostolidis, Mihir Kakara, Mark M. Painter, Rishi R. Goel, Divij Mathew, Kerry Lenzi, Ayman Rezk, Kristina R. Patterson, Diego A. Espinoza, Jessy C. Kadri, Daniel M. Markowitz, Clyde Markowitz, Ina Mexhitaj, Dina Jacobs, Allison Babb, Michael R. Betts, Eline T. Luning Prak, Daniela Weiskopf, Alba Grifoni, Kendall A. Lundgreen, Sigrid Gouma, Alessandro Sette, Paul Bates, Scott E. Hensley, Allison R. Greenplate, E. John Wherry, Rui Li, Amit Bar-Or

## Abstract

SARS-CoV-2 mRNA vaccination in healthy individuals generates effective immune protection against COVID-19. Little is known, however, about the SARS-CoV-2 mRNA vaccine-induced responses in immunosuppressed patients. We investigated induction of antigen-specific antibody, B cell and T cell responses in patients with multiple sclerosis on anti-CD20 (MS-aCD20) monotherapy following SARS-CoV-2 mRNA vaccination. Treatment with aCD20 significantly reduced Spike and RBD specific antibody and memory B cell responses in most patients, an effect that was ameliorated with longer duration from last aCD20 treatment and extent of B cell reconstitution. In contrast, all MS-aCD20 patients generated antigen-specific CD4 and CD8 T-cell responses following vaccination. However, treatment with aCD20 skewed these responses compromising circulating Tfh responses and augmenting CD8 T cell induction, while largely preserving Th1 priming. These data also revealed underlying features of coordinated immune responses following mRNA vaccination. Specifically, the MS-aCD20 patients who failed to generate anti-RBD IgG had the most severe defect in cTfh cell responses and more robust CD8 T cell responses compared to those who generated anti-RBD IgG, whose T cell responses were more similar to healthy controls. These data define the nature of SARS-CoV-2 vaccine-induced immune landscape in aCD20-treated patients, and provide insights into coordinated mRNA vaccine-induced immune responses in humans. Our findings have implications for clinical decision-making, patient education and public health policy for patients treated with aCD20 and other immunosuppressed patients.

## Introduction

Coronavirus disease 19 (COVID-19) has caused a global pandemic with profound public health and socioeconomic sequelae due to absence of protective immunity to SARS-CoV-2, the viral infectious cause of COVID-19^1, 2^. Vaccine-based strategies were rapidly developed with goals of protecting individuals and achieving herd immunity that limits transmission and subsequent infection^3^. The two mRNA vaccines that have been approved under Emergency Use Authorization (EUA) by the FDA in the US, BNT162b2 (Pfizer-BioNTech) and mRNA-1273 (Moderna), were studied in large phase-3 clinical trials and have shown high efficacy in preventing severe COVID-19 and transmissibility in healthy individuals^4, 5^. Individuals with underlying autoimmune conditions, including multiple sclerosis (MS), and those on immune-modulatory therapies were not included in these phase 3 clinical trials. As a result, the magnitude and quality of immune response to mRNA vaccination is not well characterized in these potentially vulnerable patient populations that may be at risk for higher COVID-19 associated morbidity and mortality, and more prone to infect others^6–12^.

Anti-CD20 antibody (aCD20) based B cell depleting strategies are implemented in hematologic malignancies^13^ and across a variety of autoimmune disorders^14^, including MS^15, 16^, with high rates of success. Upon antigen exposure, B cells have the ability to form memory cells or differentiate into plasmablasts and plasma cells^17^. In addition to their roles as precursors to antibody-secreting cells, B cells can function as professional antigen presenting cells, especially in the context of cognate interactions with T cells that recognize the same antigenic target^18, 19^. Depletion of B cells deprives the immune system of these B cell functions to a degree that is dependent on the depth of depletion and the sensitivity of different B cell subsets to aCD20 treatment^20–23^. As a result, vaccine-specific antibody responses are diminished in patients on aCD20 therapy^23–28^. In the case of SARS-CoV-2 mRNA vaccination, B cell depletion has been shown to result in decreased Spike-specific binding and neutralizing antibodies in patients with chronic inflammatory diseases^29^, including patients with MS^30^. Only ∼1/3 of MS patients on B cell depleting therapies had detectable antibodies ∼2-8 weeks after the second dose of mRNA vaccine^29, 30^ but the kinetics of those antibody responses, and their relationship to the level of peripheral B cell depletion and spike-specific memory B cell responses are poorly understood. Moreover, how aCD20 therapy affects other B cell dependent functions following SARS-CoV-2 mRNA vaccination is not known.

The role of B cells in T cell priming, differentiation and proliferation following an encounter with an antigen that generates a T cell dependent response remains incompletely understood, and most reports exploring this particular question are based on animal models. Specifically, some studies suggest that B cells are not required for the priming of T cell responses^31–33^ whereas other work provides evidence for B cells as antigen-presenting cells that alone, or together with dendritic cells, facilitate T-cell priming^19, 34–39^. These observations are particularly relevant in the case of SARS-CoV-2 infection and vaccination. In COVID-19, robust and long-lasting antigen-specific CD4 and CD8 T cell immunity is generated with T cell responses correlating with better outcomes in some settings^40–42^. In addition, robust CD8 T cell responses are associated with improved survival in patients with hematological malignancies and COVID-19, including patients on therapies that deplete B cells^43^. These data provide evidence that T cells may be capable of providing protective immunity and limiting severe disease in settings where antibody responses are lacking. In healthy subjects, antigen-specific CD4 T cells are generated rapidly after the first dose of SARS-CoV-2 mRNA vaccine and these responses provide the foundation for a coordinated immune response following the second vaccine dose^44^. Moreover, T cell responses have been suggested to contribute to vaccine efficacy during the early period after the first vaccine dose^44–46^. In addition, T cells are capable of recognizing mutant SARS-CoV-2 variants^47, 48^ that can partially escape humoral-based immunity. This T cell recognition of variants is likely based on broader epitope recognition including epitopes unrelated to antibody binding sites where antibody escape mutations occur^49^. Despite these data, the induction of T cell responses by mRNA vaccination in patients on B cell depleting therapies remains poorly understood. Defining the dynamics, magnitude and coordination of CD4 and CD8 T cell responses to SARS-CoV-2 mRNA vaccine in MS patients treated with aCD20 therapy should provide a better understanding of SARS-CoV-2 immunity in these patients and provide insights into the role of B cells in vaccine-induced T cell priming in humans.

In this study, we analyzed a cohort of patients with MS to evaluate the effect of aCD20 therapy on SARS-CoV-2 mRNA vaccine responses. Although most MS patients treated with aCD20 (MS-aCD20) made detectable Spike binding antibodies and 50% made detectable Receptor Binding Domain (RBD) antibodies, antibody titers were lower and delayed compared to healthy control subjects. All MS-aCD20 treated patients developed Spike-specific CD4 T cell responses, though these responses were of lower magnitude compared to healthy controls. In contrast, CD8 T cell responses were more robust in MS-aCD20 patients especially following the second vaccine dose. Finally, comparing the subsets of MS-aCD20 patients who did and did not generate anti-RBD IgG responses revealed major differences in immune response coordination, with substantial reduction in vaccine-induced cTfh responses and reciprocal increases in CD8 T-cell responses in those who failed to generate anti-RBD antibodies. Thus, these data demonstrate the efficient induction of T cell immunity in MS-aCD20 patients following SARS-CoV-2 mRNA vaccination even in the absence of antibody induction. Moreover, these studies provide insights into the role of B cells and humoral immunity in vaccine-induced T cell responses and shed light on immune mechanisms that accompany aCD20 therapy based on differential responses to vaccination.

## Results

### Impact of aCD20 therapy on SARS-CoV-2 mRNA vaccine-induced antibody responses

Whereas SARS-CoV-2 mRNA-based vaccination has been successful in generating robust B cell and T cell mediated vaccine responses in healthy individuals^44, 45, 47, 48, 50, 51^, less is known about responses to these vaccines in patients with autoimmune conditions, particularly those receiving immunosuppressive or immune cell-depleting agents^29, 52^. To examine the impact of anti-CD20 (aCD20) monoclonal antibody therapy on responses to SARS-CoV-2 mRNA vaccination, we recruited 20 patients with multiple sclerosis (MS) treated with aCD20 (MS-aCD20) and compared their vaccine-induced immune responses to 10 healthy control (HC) subjects within the University of Pennsylvania Health System (**Extended Data Table 1**). Plasma and peripheral blood mononuclear cell (PBMC) samples were analyzed at 5 timepoints (Figure 1A): prior to the 1^st^ vaccine dose (timepoint 1, baseline), 10-12 days following the 1^st^ vaccine dose (timepoint 2), prior to the 2^nd^ vaccine dose (timepoint 3), 10-12 days following the 2^nd^ vaccine dose (timepoint 4) and 25-30 days following the 2^nd^ vaccine dose (timepoint 5). We evaluated the kinetics of humoral and cellular vaccine responses from induction to early memory.

**Figure 1.**
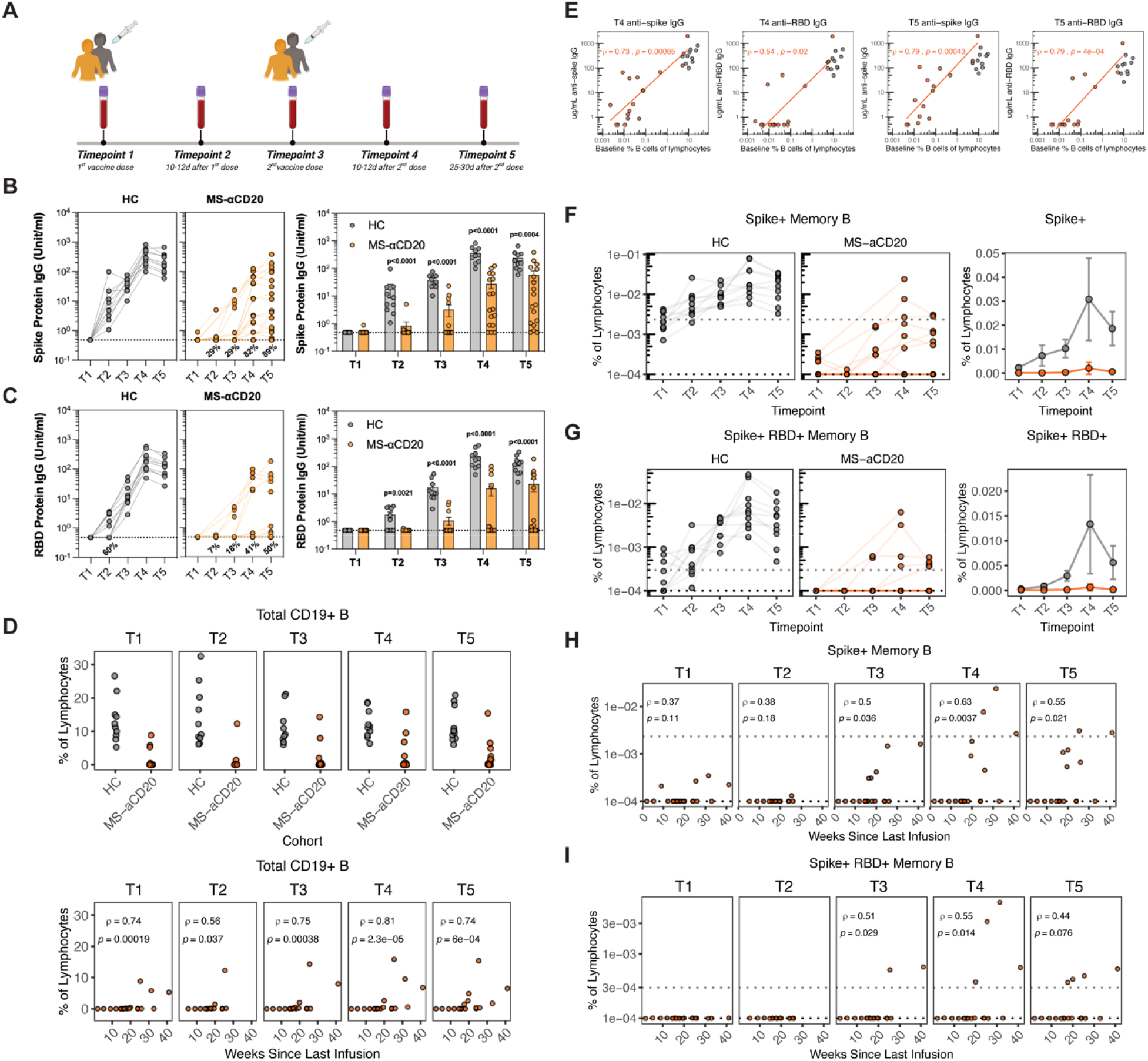
Decreased humoral responses following SARS-CoV-2 mRNA vaccination in MS-aCD20 patients. **A)** The longitudinal study design, vaccine administration scheme and timepoints collected following SARS-CoV-2 mRNA vaccination for the HC and MS-aCD20 patient cohorts. **B-C)** Anti-Spike IgG **(B)** and anti-RBD IgG **(C)** for all timepoints collected (T1-T5) were measured in the HC and MS-aCD20 patients. Results are shown as units/mL, using unpaired, two-tailed, non-parametric Wilcoxon testing for analysis. **D)** Frequency of CD19^+^ B cells as a percentage of total lymphocytes in healthy controls (HC) and MS-aCD20 patients **(top)**. Correlation between frequency of total CD19^+^ B cells and weeks since last infusion of anti-CD20 **(bottom)**. Correlations were calculated using non-parametric Spearman rank correlation. **E)** Correlations between the frequency of baseline (T1) percentage of B cells of all lymphocytes and the levels of anti-spike IgG at T4 and T5 **(left)** or anti-RBD IgG at T4 and T5 **(right)** following vaccination. Gray points represent HCs, orange points represent MS-aCD20 patients. Only MS-aCD20 patients were considered for the correlations, with HCs as a visual reference. Associations were calculated using Spearman rank correlation and are shown with Pearson trend lines for visualization. **F-G)** Frequency of Spike^+^ **(F)** and Spike^+^RBD^+^ **(G)** memory B cells over time in vaccinated individuals. Data are represented as frequency of antigen-specific cells in the total lymphocyte compartment (left panels log scale, right panel linear scale). **H-I)** Correlation between frequency of Spike^+^ **(H)** and Spike^+^RBD^+^ **(I)** memory B and weeks since last infusion of anti-CD20. Correlations were calculated using non-parametric Spearman rank correlation.

Serological responses to SARS-CoV-2 Spike and RBD proteins are evidence of productive immune responses and vaccine efficacy^40, 53–55^. To interrogate serological outcomes in this MS-aCD20 cohort, we quantified anti-Spike and anti-RBD IgG following the first and second doses of mRNA vaccination (Figure 1B-C). All HC subjects generated both anti-Spike and anti-RBD IgG following the first dose of mRNA vaccine and the level of antibody increased further after the second dose (Figure 1B-C and **Extended Data Table 2**), as reported previously^51^. In contrast, these responses were considerably more variable in the MS-aCD20 patients, with 85% developing detectable anti-Spike IgG and only 50% mounting detectable anti-RBD IgG responses by timepoint 5 (Figure 1B-C and **Extended Data Table 2**). Among those MS-aCD20 patients with detectable IgG, the magnitude of response was generally lower, and the kinetics of the IgG response were delayed compared to the HC group. These findings extend the prior observations^29, 30^ that antibody responses to SARS-CoV-2 mRNA vaccine are substantially attenuated in MS patients on aCD20 therapy. Our data indicate that many of these patients can still mount detectable antibody responses, albeit with decreased magnitude, delayed kinetics and considerable heterogeneity across patients.

Since a major reason for the substantially altered antibody responses in MS-aCD20 patients was likely B cell depletion, we considered whether the heterogeneity in antibody responses (Figure 1B and C) was related to the duration between the vaccination and the last aCD20 infusion. There were trends towards increased serologic responses to both Spike (**Extended Data Figure 1A**) and RBD (**Extended Data Figure 1B**) as the duration from the last aCD20 infusion increased. To further test this idea, we quantified total CD19^+^ B cell numbers in the circulation of the MS-aCD20 and HC groups (**Extended Data Figure 1C).** Although most MS-aCD20 patients had no detectable B cells, small circulating B cell populations were detectable in some patients, and there was a clear relationship between time since last aCD20 infusion and the extent of B cell reconstitution at all timepoints examined (Figure 1D). MS-aCD20 patients with higher percentages of circulating B cells prior to the vaccine (T1) had more robust anti-Spike and anti-RBD IgG responses at T4 and T5 (Figure 1E). The small number of MS-aCD20 patients who had circulating B cell frequencies comparable to the healthy control group achieved equivalent antibody titers following vaccination (Figure 1E), suggesting that B cells repopulating the peripheral pool after aCD20 infusion are functionally competent. Thus, when the circulating B cell pool is repopulated in patients, as seen with increased duration from their last aCD20 administration, vaccine-induced antibody responses can approach levels observed in healthy control subjects. Moreover, the data above demonstrate that even patients who are lacking or are severely deficient in detectable B cells in their circulation can still mount some antibody responses to mRNA vaccines, although with substantially reduced magnitude.

### aCD20 effects on vaccine-induced antigen-specific memory B cells

Antigen-specific memory B cells represent a key feature of long-term immunity^56^. These cells are able to respond rapidly to subsequent infections, generate new antibody secreting cells and initiate new germinal center reactions where antibody can further improve qualitatively through somatic hypermutation and affinity maturation^51, 56–59^. Using biotinylated Spike and RBD B cell probes, we recently investigated the antigen-specific memory B cell responses following SARS-CoV-2 mRNA vaccination in healthy individuals^51^. We employed this same strategy to define the magnitude and kinetics of the memory B cell response generated in MS-aCD20 patients following SARS-CoV-2 mRNA vaccination. While circulating memory B cells specific for both Spike (**Extended Data Figure 1D** and Figure 1F) and RBD (**Extended Data Figure 1D** and Figure 1G) were readily induced in all HCs, Spike-specific memory B cells were detected in only a subset of MS-aCD20 patients, where their frequencies were also substantially diminished (Figure 1F) at all timepoints (**Extended Data Table 3**). Similarly, only a minority of MS-aCD20 patients had detectable RBD-specific memory B cells in the circulation at any timepoint (Figure 1G and **Extended Data Table 3**). Finally, in agreement with the data above on antibody responses and total B cells, there was a strong correlation between the ability to detect antigen-specific memory B cells and a longer period of time since the last aCD20 treatment (Figure 1H-I). Thus, induction of antigen-specific memory B cell responses following SARS-CoV-2 mRNA vaccination was compromised in MS-aCD20 patients compared to HCs and this impairment was more pronounced in patients who were immunized in closer proximity to their last aCD20 infusion. There were substantially more patients with detectable antibody responses (88.9%) than patients with detectable circulating memory B cells (30%) to the Spike antigen. While this discrepancy may reflect limits of detection of the antigen-specific memory B cell assay, an alternate explanation is that early repopulation of functional B cells within lymphoid tissues enables productive antibody responses even prior to re-emergence of detectable total B cells or antigen-specific memory B cells in the circulation.

### aCD20 impact on vaccine-induced CD4 T cell responses

Whereas the induction of B cell responses and antibody responses is a major goal of vaccination against SARS-CoV-2 given the key role of antibodies in protecting from infection, B cells are also central to the effective priming, differentiation, proliferation and maintenance of T cell responses following immunization or infection. Yet the impact of B-cell depletion due to aCD20 treatment on T cell responses to SARS-CoV-2 mRNA vaccination is unclear. Thus, to interrogate the effect of aCD20 treatment on human vaccine-induced T cell responses, we initially implemented high-dimensional flow cytometric analysis of circulating T cell populations in the MS-aCD20 patients and HCs following SARS-CoV-2 mRNA vaccination. Specifically, we applied an unbiased approach using opt-SNE^60^ dimensionality reduction followed by FlowSOM clustering^61^ (**Extended Data Figure 3**). Examining the total CD4^+^ T cell landscape over time revealed dynamic changes following mRNA vaccination (**Extended Data Figure 3A**). The total landscape could be mapped with key markers (**Extended Data Figure 3B**) and metaclusters corresponding to distinct subpopulations of CD4 T cells (**Extended Data Figure 3C-D**). We identified a group of small metaclusters (metaclusters 9-14) that expanded following the first vaccine dose in HC subjects and expressed high Ki67, CD38, ICOS and HLA-DR, consistent with induction of activated T cells responding to vaccination. This group of metaclusters showed less dynamic change in the MS-aCD20 group with more subtle induction at T2 and T4. No differences were observed in the abundance of these metaclusters between the MS-aCD20 and HC groups at either T2 or T4 (**Extended Data Figure 3E**). This high-dimensional single cell cytometry-based analysis revealed global changes in responding CD4 T cell populations in both MS-aCD20 and HC groups. We next wanted to gain deeper insights into the CD4 T cell subpopulations induced by vaccination, and identify potential differences between the MS-aCD20 and HC groups.

Human vaccination induces populations of Ki67^+^CD38^+^ CD4 and CD8 T cells ∼1-2 weeks post immunization and this activated, proliferating subset, contains antigen-specific T cells^62–65^. We therefore initially examined the induction and dynamics of Ki67^+^CD38^+^ CD4 T cells (gating strategy in **Extended Data Figure 2**). Consistent with previous reports^44^, a population of Ki67^+^CD38^+^ CD4 T cells was robustly induced after the first vaccine dose in HCs, peaking at T2 and then returning to baseline (Figure 2A). In contrast, although MS-aCD20 patients had similar frequencies of activated CD4 T cells at baseline, the induction of Ki67^+^CD38^+^ CD4 T cells following vaccination was reduced compared to the HC subjects at T2 with no increase noted after the second dose and the frequency of activated CD4 T cells in MS-aCD20 patients remained lower than HC through T5 (Figure 2A). We next applied dimensionality reduction and FlowSOM-based clustering only on these activated Ki67^+^CD38^+^ CD4 T cells (Figure 2B-F). Comparison of MS-aCD20 patients to HCs revealed landscape differences independent of vaccination or timepoint (Figure 2B). However, there were also clear patterns of vaccine-induced change in subpopulations of Ki67^+^CD38^+^ CD4 T cells. There were areas of more intense Ki67 or CD38 expression, as well as areas of cells that expressed FoxP3, CTLA-4, CXCR5, CXCR3, CCR6, Tbet and other activation markers corresponding to distinct subpopulations of activated CD4 T cells (Figure 2C-E and **Extended Data Figure 4**). Additional analysis identified metaclusters with clear enrichment following vaccination. Specifically, metacluster 1 increased at T2 and metacluster 7 increased at both T2 and T4 (Figure 2F). Metacluster 1 was composed of highly activated Ki67^++^ICOS^++^CXCR3^+^T-bet^mid^ CD4 T cells of the CM/EM1 memory phenotype (CM/EM1 Th1 cells). Metacluster 7 represented a combination of discrete CCR6^+^T-bet^-^ or CXCR3^+^T-bet^mid^ CM/EM1 CD4 T cells with high ICOS (CM/EM1 Th17-like and Th1-like cells). The dynamic changes in these two metaclusters following vaccination were similar between the MS-aCD20 and HC groups (Figure 2F). We next sought to understand the response of circulating T follicular helper (cTfh) cells given the role of Tfh cells in supporting antigen-specific B cell responses. Metacluster 3 was an activated (CD38^+^ ICOS^+^ HLA-DR^+^), proliferating (Ki67^+^) subpopulation with high expression of CXCR5 and PD1 (Figure 2E), corresponding to activated cTfh cells. This metacluster was similarly induced after the first vaccine dose for both MS-aCD20 patients and HC (Figure 2F; **Extended Data Figure 4B**). However, following the second vaccine dose and through the early memory timepoint (T5), metacluster 3 decreased in proportion (Figure 2F) and contracted (**Extended Data Figure 4B**) in MS-aCD20 patients compared to HC. Thus, this analysis identified subpopulations of CD4 T cells that responded similarly to vaccination when comparing MS-aCD20 patients to HC subjects (e.g. subsets of activated Th1 cells) as well as a population of cTfh cells that had similar initial induction in the two cohorts, but poor maintenance at later timepoints in the MS-aCD20 patients.

**Figure 2.**
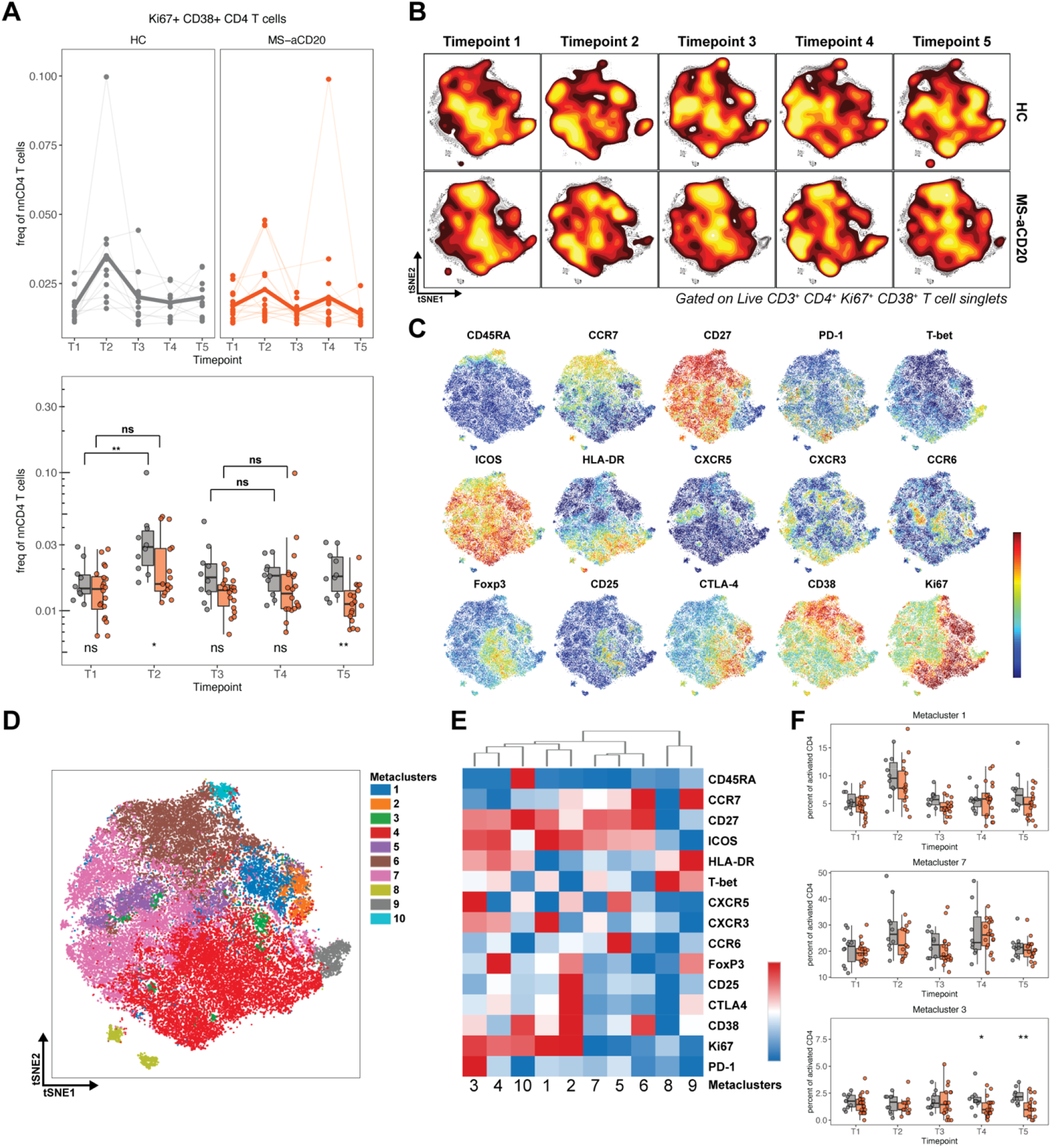
SARS-CoV-2 mRNA vaccination results in altered CD4 T cell activation in MS-aCD20 patients. **A)** The frequency of activated Ki67^+^CD38^+^ CD4 T cells of total non-naïve CD4 T cells. Top: individual subjects (points) and the mean (thicker line) are shown for each group. Bottom: Tukey boxplots for each timepoint and group are depicted. Unpaired Wilcoxon test was used to compare the two groups at each timepoint (shown under the boxplots) or the groups between the timepoints indicated (shown on top of the boxplots). **B)** Opt-SNE projections of concatenated cytometry data for activated Ki67^+^CD38^+^ CD4 T cells for each timepoint and group combination are shown. **C)** Surface expression intensity of the markers indicated on the opt-SNE 2D-map generated with all samples in B. **D)** FlowSOM metaclusters were created using activated Ki67^+^CD38^+^ CD4 T cells concatenated from all samples and projected to the opt-SNE map. **E)** Surface expression intensity heatmap of the markers indicated for each of the 10 FlowSOM metaclusters in D. **F)** The abundance of metaclusters 1, 7 and 3 as percentage of activated Ki67^+^CD38^+^ CD4 T cells is shown for HC (grey) and MS-aCD20 (orange) groups; unpaired Wilcoxon test *p* values are shown when *p* < 0.05 between groups.

To better understand whether these vaccine-induced changes in CD4 T cells reflected antigen-specific CD4 T cell responses, we performed peptide-dependent activation-induced marker (AIM) assays. Specifically, we stimulated PBMCs with a peptide megapool containing SARS-CoV-2 Spike epitopes optimized for presentation by MHC-II (CD4-S), as previously described^44, 66, 67^. AIM^+^ CD4^+^ T cells were defined by co-expression of CD200 and CD40L (Figure 3A and **Extended Data Figure 5A**). We first confirmed that the absence of B cells during the AIM peptide stimulation assay did not impact the readout of AIM^+^ T cells (**Extended Data Figure 5B**) allowing direct comparisons between the MS-aCD20 and HC PBMCs with this assay. Following the first dose of the SARS-CoV-2 mRNA vaccine, both MS-aCD20 and HC groups demonstrated robust increases in their AIM^+^ CD4 T cells indicating efficient CD4 T cell priming (Figure 3B). HC subjects retained high AIM^+^ CD4 T-cell frequencies at all subsequent timepoints with a trend towards an additional increase following the second vaccine dose (Figure 3B), an effect we previously reported in a larger cohort of healthy individuals^44^. Compared to HCs, AIM^+^ CD4 T cell responses in the MS-aCD20 patients were modestly attenuated at T3 but nonetheless efficiently boosted by the second vaccine dose, and again somewhat diminished compared to HCs at T5 (Figure 3B). To further assess memory T cell subsets at the antigen-specific level, we subdivided the AIM^+^ CD4 T cells into CM, three different subpopulations of effector memory T cells (EM1, EM2, EM3) and EM T cells re-expressing CD45RA (EMRA) (Figure 3C and **Extended Data Figure 5C**). There were no major differences in the distribution of AIM^+^ CD4 T cells among these memory T cell subsets between MS-aCD20 patients and HCs, with the majority of AIM^+^ CD4 T cells mapping to the CM and EM1 subsets (Figure 4D) in both groups. Similarly, we used CXCR5, CXCR3 and CCR6 (Figure 3E and **Extended Data Figure 5D**) to examine CD4 T helper subsets among antigen-specific AIM^+^ CD4 T cells in the MS-aCD20 and HC cohorts. Although the distribution was largely similar between the cohorts, there was a trend towards a lower frequency of cTfh cells among the total AIM^+^ responding CD4 T cells in the MS-aCD20 group (Figure 3F and **Extended Data Figure 5E**). Thus, although there were some modest reductions at particular timepoints in the MS-aCD20 patients compared to HCs, these data indicate that MS-aCD20 patients are capable of generating robust antigen-specific CD4 T cell responses to both vaccine doses despite attenuated antibody responses.

**Figure 3.**
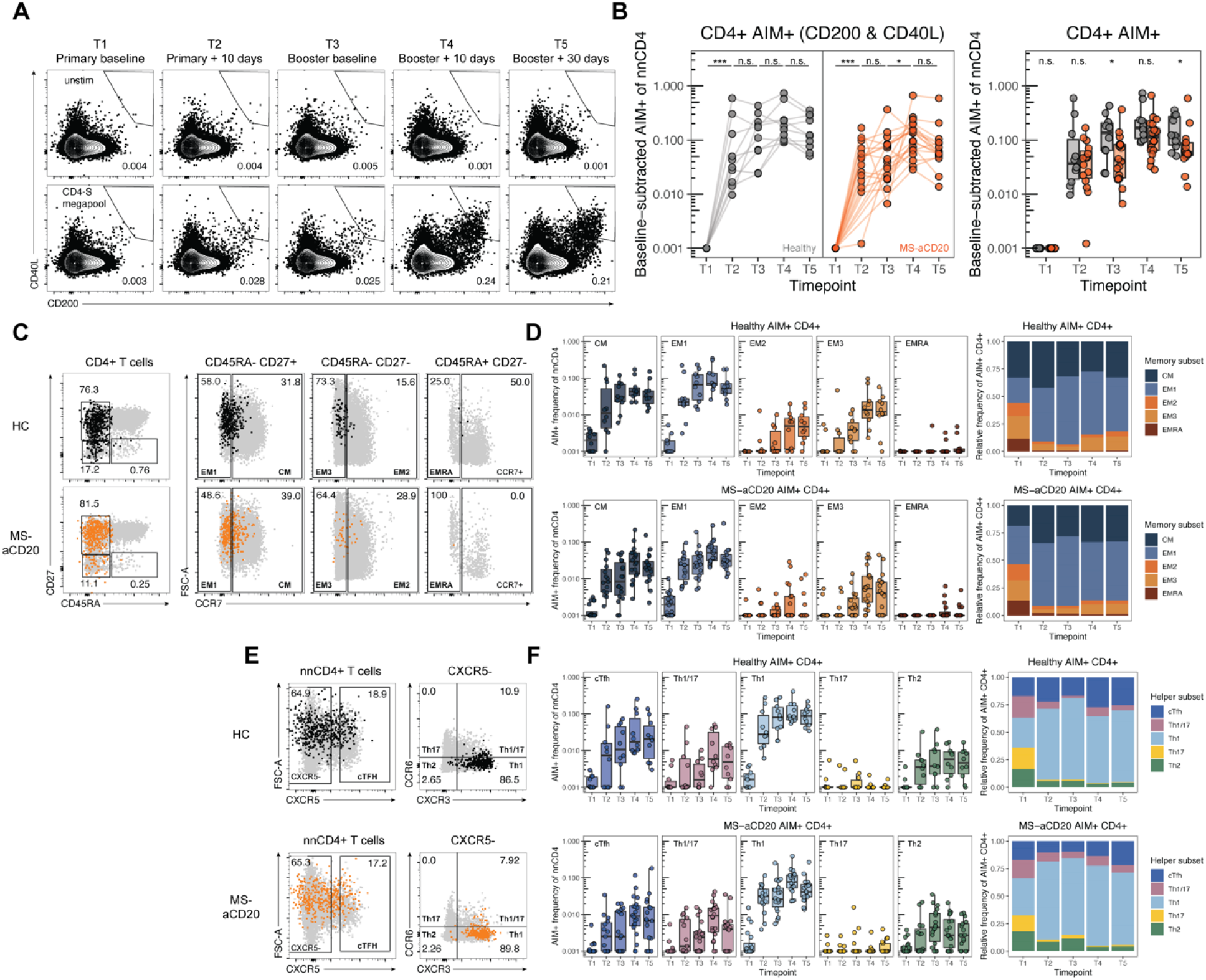
SARS-CoV-2 mRNA vaccination elicits near-normal antigen-specific CD4 T cell responses in MS-aCD20 patients. **A)** Representative flow cytometry plots for quantification of AIM^+^ CD4 T cells. Numbers represent the percentage of total non-naïve CD4 T cells that are AIM^+^. Top row is unstimulated, bottom row is stimulated with the CD4-S megapool. **B)** Summary data of AIM^+^ CD4 T cell frequency following vaccination. Values represent the background-subtracted frequency of AIM^+^ non-naïve CD4 T cells above paired background-subtracted baseline frequencies. Lines connect individual donors sampled longitudinally. Statistics were calculated using unpaired Wilcoxon test. Gray indicates HCs, orange indicates patients with MS receiving anti-CD20 therapy. **C)** Representative plots demonstrating the identification of the indicated memory T cell subsets from AIM^+^ CD4 T cells. Black or orange events depict AIM^+^ cells from HC or MS-aCD20 subjects, respectively. Gray events depict total CD4 T cells from the same donor. Numbers indicate the frequency of AIM^+^ cells within each gate. **D)** Frequency of memory T cell subsets in AIM^+^ CD4 T cells. Top panels depict healthy controls. Bottom panels depict subjects with MS receiving anti-CD20 therapy. Left panels depict the background-subtracted percent of non-naïve T cells that are AIM^+^ cells of each subset. Right panels depict the relative frequency of each memory T cell subset in the background-subtracted AIM^+^ population. CM = CD45RA^-^CD27^+^CCR7^+^, EM1 = CD45RA^-^ CD27^+^CCR7^-^, EM2 = CD45RA^-^CD27^-^CCR7^+^, EM3 = CD45RA^-^CD27^-^CCR7^-^, EMRA = CD45RA^+^CD27^-^CCR7^-^. **E)** Representative flow cytometric plots depicting the gating of AIM^+^ CD4 T cells to identify the indicated helper subsets as in (C). **F)** Frequency of T helper subsets in AIM^+^ CD4 T cells as in (D). cTfh = CXCR5^+^ of non-naïve CD4 T cells, Th1 = CXCR5^-^CXCR3^+^CCR6^-^, Th2 = CXCR5^-^CXCR3^-^CCR6^-^, Th17 = CXCR5^-^CXCR3^-^ CCR6^+^, Th1/17 = CXCR5^-^CXCR3^+^CCR6^+^.

**Figure 4.**
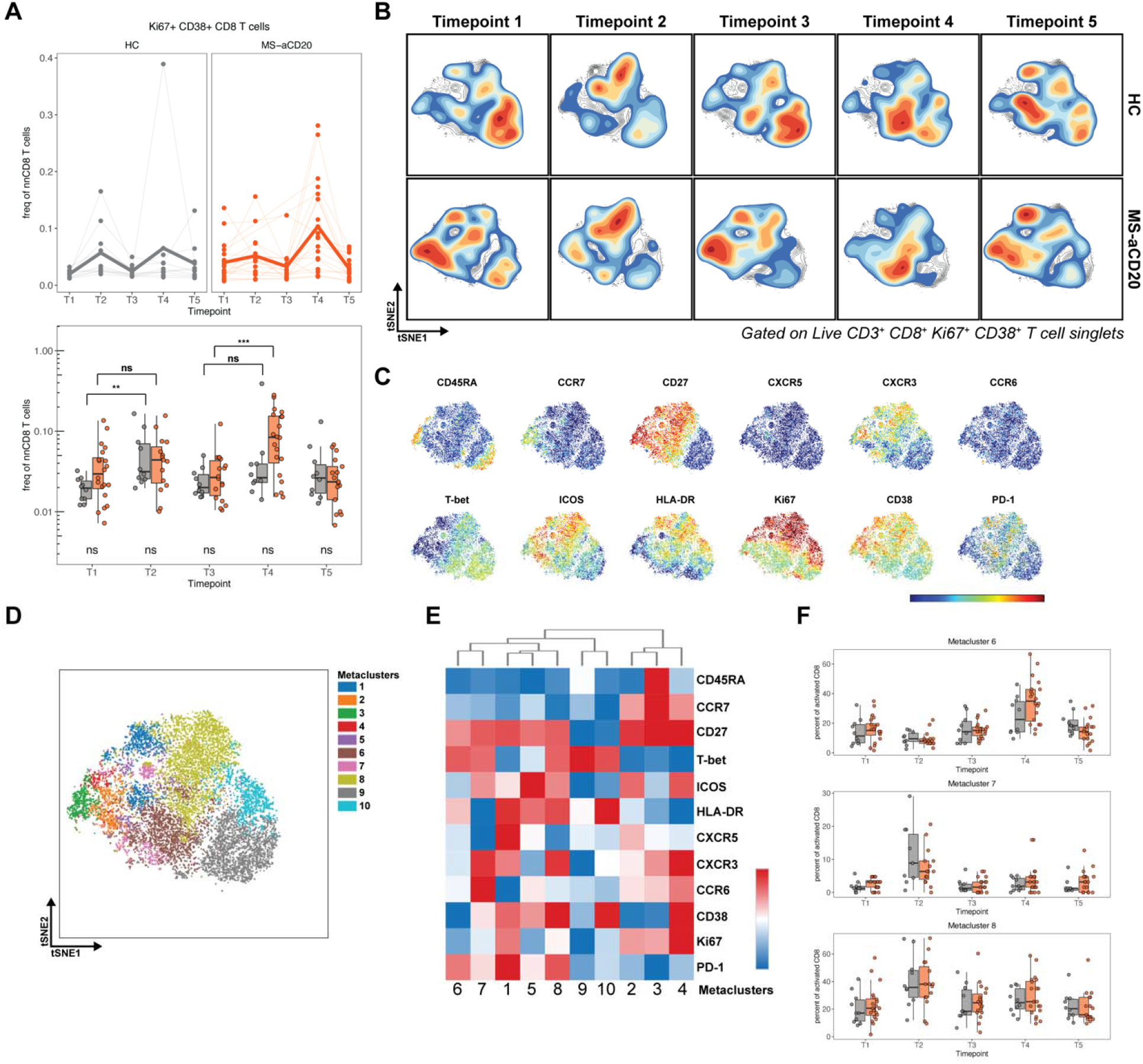
SARS-CoV-2 mRNA vaccination results in robust CD8 T cell activation in HCs and MS-aCD20 patients. **A)** The frequency of activated Ki67^+^CD38^+^ CD8 T cells of total non-naïve CD8 T cells is depicted. Top: individual subjects (points) and the mean (thicker line) are shown for each group. Bottom: Tukey boxplots for each timepoint and group are depicted. Unpaired Wilcoxon test was used to compare the two groups at each timepoint (shown under the boxplots) or the groups between timepoints indicated (shown on top of the boxplots). **B)** Opt-SNE projections of concatenated cytometry data for activated Ki67^+^CD38^+^ CD8 T cells for each timepoint and group combination are shown. **C)** Surface expression intensity of the markers indicated on the opt-SNE 2D-map generated with all samples in B. **D)** FlowSOM metaclusters were created using activated Ki67^+^CD38^+^ CD8 T cells concatenated from all samples and projected onto the opt-SNE map. **E)** Surface expression intensity heatmap of the markers indicated for each of the 10 FlowSOM metaclusters in D. **F)** The abundance of metaclusters 6, 7 and 8 as percentage of activated Ki67^+^CD38^+^ CD8 T cells is shown for HC (grey) and MS-aCD20 (orange) groups; unpaired Wilcoxon test *p* values are shown when *p* < 0.05 between groups.

### aCD20 impact on vaccine-induced CD8 T cell responses

We next examined CD8 T cell responses following vaccination in MS-aCD20 patients and HCs. We first assessed activated Ki67^+^CD38^+^ CD8 T cells (Figure 4) using the approaches outlined above for CD4 T cells (Figure 2). Both doses of vaccination elicited increases in activated CD8 T cells in HCs and MS-aCD20 patients (Figure 4A). Activated CD8 T cells moderately expanded after the first vaccine dose in both cohorts, though the magnitude of increase was more robust for HCs (Figure 4A), possibly due to higher prevaccination (T1) CD8 T cell activation in the MS-aCD20 group. The MS-aCD20 patients, however, generated a considerably stronger response to the second vaccine dose than the HC group, suggesting more robust induction of CD8 T cell responses in the B cell depleted state. We next applied the metaclustering approach described above for CD4 T cells to interrogate the phenotype of the vaccine-responding activated Ki67^+^CD38^+^ CD8 T cells (Figure 4B-F and **Extended Data Figure 6**). The opt-SNE landscape map of activated CD8 T cells revealed differences between MS-aCD20 patients and HCs prior to the vaccine, including an abundance of CD27^+^ICOS^+^CD38^+^ CD8 T cells largely lacking T-bet in MS-aCD20 patients in contrast to CD27^-^T-bet^+^ CD8 T cells in HCs (Figure 4B). However, the activated CD8 T cell populations in both MS-aCD20 patients and HC subjects reoriented following the two vaccine doses, such that they occupied similar opt-SNE space (Figure 4B). Metaclusters were generated to better define these vaccine-induced changes (Figure 4D-F and **Extended Data Figure 6**). Specifically, metacluster 7 and 8 were the main vaccine-responding CD8 T cell populations in both groups following the first vaccine dose (Figure 4B and Figure 4F). Metacluster 6 was the predominant population enriched after the second vaccine dose in both groups (Figure 4B and Figure 4F). All three metaclusters were EM1 populations that expressed T-bet and PD-1, although the metaclusters responding to the primary vaccination had higher levels of CXCR3, ICOS and CD38 (Figure 4E). Thus, these high dimensional cytometry data indicated that vaccine-induced activated CD8 T cell responses were more robust in MS-aCD20 patients compared to HC subjects after the second vaccine dose. Moreover, SARS-CoV-2 vaccination appeared to reorganize the distinct baseline landscapes of CD8 T cell activation in MS-aCD20 and HC subjects, reflecting similar vaccine-induced activation profiles.

We next performed AIM assays on CD8 T cells stimulated with a CD8-E peptide megapool as described^66, 67^ (Figure 5) to examine antigen-specific CD8 T cell responses. As previously reported^44^, AIM^+^ CD8 T cell responses were detected in a subset of HC subjects following the first vaccine dose, with more individuals responding following the second vaccine dose (Figure 5A-B). A similar pattern was seen in the MS-aCD20 patients. However, following the second vaccine dose (T4), a significantly greater expansion of antigen-specific CD8 T cells was noted in the MS-aCD20 patients compared to HCs, a difference that persisted at T5. This expansion was dominated by EM1 cells (Figure 5C-D) consistent with the observations above using activation markers (Figure 4F). Of note, both groups had equivalent frequencies of total memory subsets that were largely unchanged during vaccination (**Extended Data Figure 7**). Thus, although the overall distribution of memory CD8 T cell subsets was similar, SARS-CoV-2 mRNA vaccination induced a stronger antigen-specific CD8 T cell response in MS-aCD20 patients compared to HCs, in particular following the second dose of vaccine.

**Figure 5.**
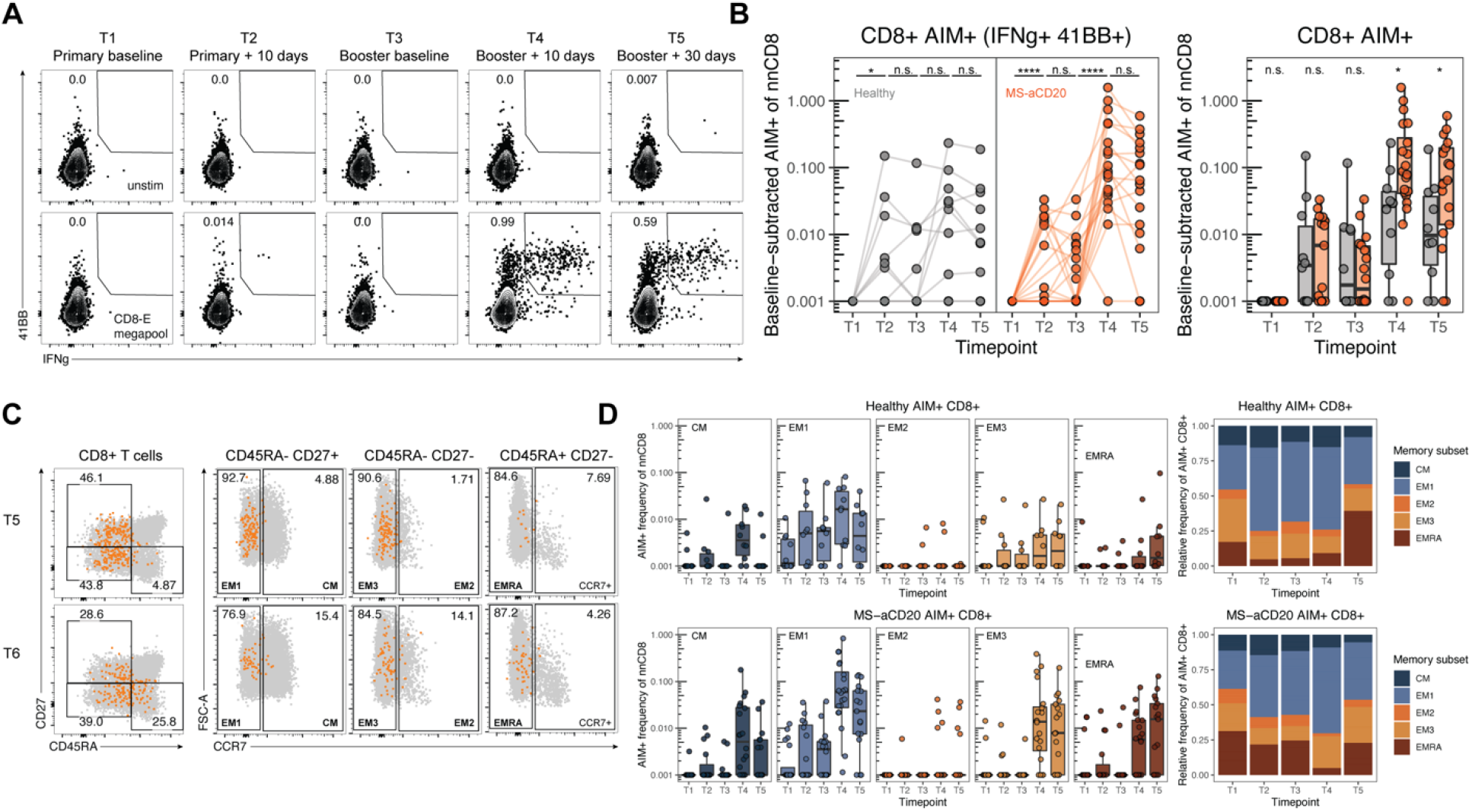
Enhanced antigen-specific CD8 T cell responses following mRNA vaccination in MS-aCD20 patients. **A)** Representative flow cytometry plots for quantifying AIM^+^ CD8 T cells. Numbers represent the percentage of total non-naïve CD8 T cells that are AIM^+^. Top row is unstimulated, bottom row is stimulated with the CD8-E megapool. **B)** Summary data of AIM^+^ CD8 T cell frequency following vaccination. Values represent the background-subtracted frequency of AIM^+^ non-naïve CD4 T cells above paired background-subtracted baseline frequencies. Lines connect individual donors sampled longitudinally. Statistics were calculated using unpaired Wilcoxon test. Gray indicates HCs, orange indicates patients with MS receiving anti-CD20 therapy. **C)** Representative flow cytometric plots depicting identification of the indicated memory T cell subsets from AIM^+^ CD8 T cells. Orange events depict AIM^+^ cells from MS-aCD20 subjects. Gray events depict total CD4 T cells from the same donor. Numbers indicate the frequency of AIM^+^ cells within each gate. **D)** Frequency of memory T cell subsets in AIM^+^ CD8 T cells. Top panels depict healthy controls. Bottom panels depict subjects with MS receiving anti-CD20 therapy. Left panels depict the background-subtracted percent of non-naïve T cells that are AIM^+^ cells of each subset. Right panels depict the relative frequency of each memory T cell subset in the background-subtracted AIM^+^ population. CM = CD45RA^-^CD27^+^CCR7^+^, EM1 = CD45RA^-^CD27^+^CCR7^-^, EM2 = CD45RA^-^CD27^-^CCR7^+^, EM3 = CD45RA^-^CD27^-^CCR7^-^, EMRA = CD45RA^+^CD27^-^CCR7^-^.

### Subsets of MS-aCD20 patients with distinct coordination of vaccine-induced immune responses

In previous work, we demonstrated that different features of the adaptive immune response were highly coordinated following SARS-CoV-2 mRNA vaccination^44^. Thus, we next wanted to examine how variation in the extent of B cell depletion and the associated impact on both B cell and humoral immune responses in the MS-aCD20 cohort might impact other features of the vaccine-induced immune response. First, comparing antigen-specific measures across timepoints T2, T4 and T5 revealed a strong correlation between humoral and cTfh responses (Figure 6A-B). This correlation was evident earlier and to a stronger extent in the MS-aCD20 group, possibly due to the larger variance of these coordinated features within the B cell depleted cohort. In contrast, AIM^+^ CD8 T cells showed a strong negative correlation with humoral immune features at T5 in the MS-aCD20 group (Figure 6A and 7C). AIM^+^ Th1 cells were also no longer positively associated with some features of humoral immunity as observed in HCs (Figure 6A). These findings prompted us to separate the MS-aCD20 patients into those who made a detectable RBD-specific IgG response (RBD Ab+) and those who did not (RBD Ab-) and then to investigate other potential immune differences between these two subgroups of MS-aCD20 patients. Figure 6D shows the opt-SNE projection of Ki67^+^CD38^+^ CD4 T cells for the three groups: HC, MS-aCD20 RBD Ab+ and MS-aCD20 RBD Ab-. The landscape of Ki67^+^CD38^+^ CD4 T cells from RBD Ab+ MS-aCD20 patients was similar to that of HC and both RBD Ab+ MS-aCD20 and HC displayed some overlapping temporal features of change during the course of vaccination. In contrast, the RBD Ab-MS-aCD20 group displayed a distinct opt-SNE projection of Ki67^+^CD38^+^ CD4 T cells that was notable for minimal vaccine-induced changes compared to both other groups. To quantify these differences, we used the Earth Mover’s Distance (EMD) metric for all pair-wise comparisons that calculates similarities between the probability distributions within the opt-SNE maps^68, 69^. EMD revealed similarity in the overall landscape of activated CD4 T cells between HC and RBD Ab+ MS-aCD20 patients, whereas the RBD Ab-MS-aCD20 group was highly dissimilar to the other two groups at all the timepoints examined (**Extended Data Figure 8A**). In contrast to activated CD4 T cells, vaccine-induced changes in the activated CD8 T cell compartment after the first dose (T2) were more similar in RBD Ab+ and Ab-MS-aCD20 groups, both of which resembled the HC responses (Figure 6E). These findings were confirmed with our EMD analysis at T2 (**Extended data Figure 8B**). Following the second vaccine dose (T4), however, the RBD Ab+ MS-aCD20 group was different from both the HC and RBD-MS-aCD20 groups (Figure 6E and **Extended data Figure 8B**) due to the larger presence of metacluster 8 (see Figure 4 for CD8 metacluster annotation). Taken together, these data show that, in the absence of a functional humoral response using anti-RBD IgG as a proxy, the defects identified in vaccine-induced responses of activated CD4 T cells in the MS-aCD20 patients were amplified. In contrast, vaccine-induced CD8 T cell responses were more similar in MS-aCD20 patients when compared to HCs with less impact of anti-RBD IgG status.

**Figure 6.**
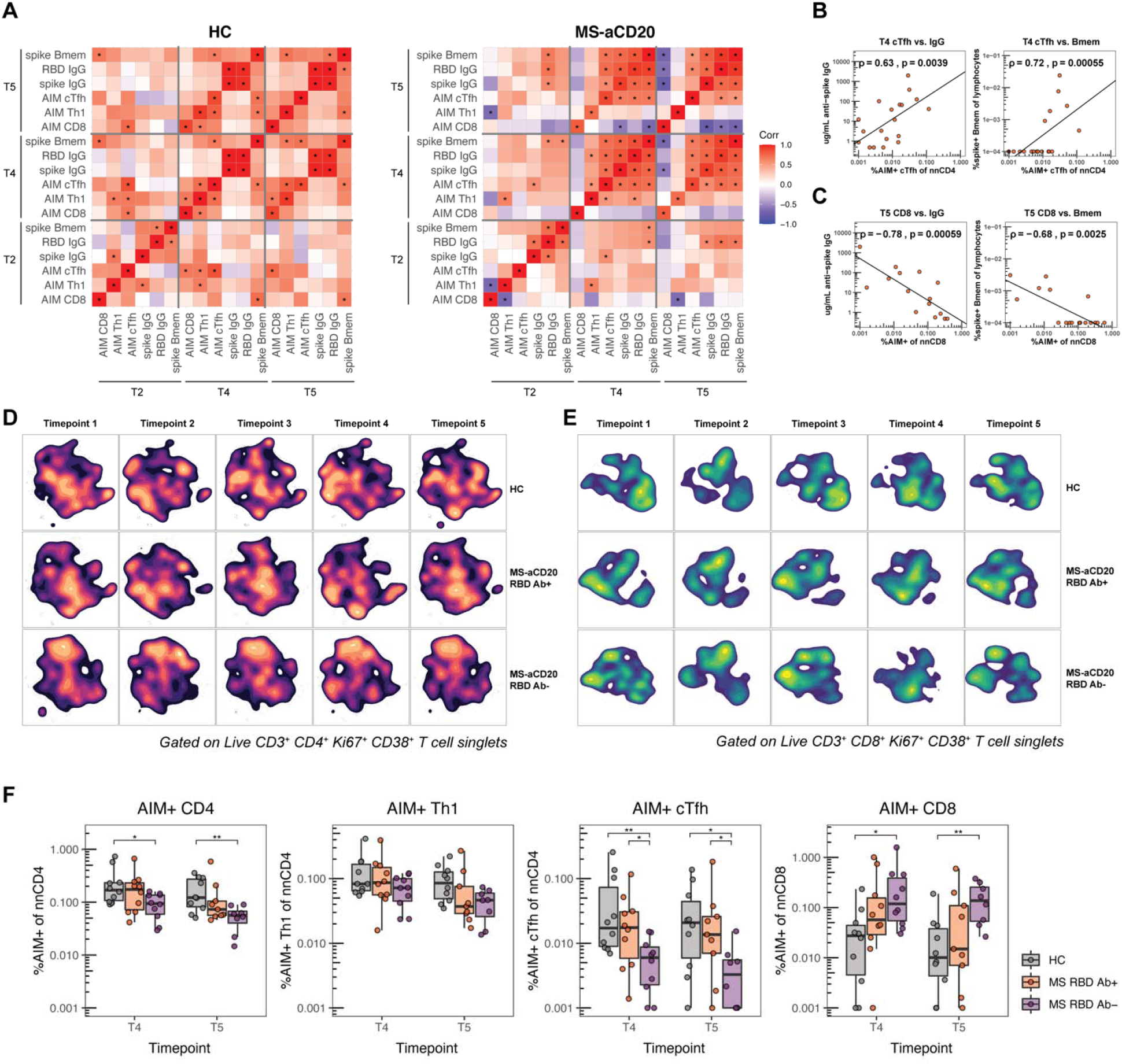
MS-aCD20 patients with no detectable anti-RBD IgG demonstrate the highest level of immune discoordination following SARS-CoV-2 mRNA vaccination. **A)** Correlations of antigen-specific features at timepoints T2, T4 and T5 in HC (left) and MS-aCD20 subjects (right). Associations were calculated using Spearman rank correlation; * indicates *p* < 0.05. **B-C)** Correlations between the frequency of T4 AIM^+^ cTfh cells and T4 anti-spike IgG (left) or spike-specific memory B cells (right) **(B)** and between the frequency of T5 AIM^+^ CD8 T cells and T5 anti-spike IgG (left) or spike-specific memory B cells (right) **(C)**. Only patients receiving anti-CD20 therapy were considered for these correlations. Associations were calculated using Spearman rank correlation and are shown with Pearson trend lines for visualization. **D-E)** Opt-SNE projections of concatenated cytometry data for activated Ki67^+^CD38^+^ CD4 **(D)** and activated Ki67^+^CD38^+^ CD8 **(E)** T cells for each timepoint and group combination are shown. Groups: healthy control (HC), MS-aCD20 with anti-RBD IgG + at any timepoint (MS-aCD20 RBD Ab+) and MS-aCD20 with anti-RBD IgG -(MS-aCD20 RBD Ab-) at all timepoints examined. **F)** AIM^+^ frequencies of the indicated T cell populations following mRNA vaccination at T4 and T5. Values represent the background-subtracted frequency of AIM^+^ non-naïve T cells above paired baseline frequencies for HC (grey) and MS RBD Ab+ (orange) and MS RBD Ab-(purple) groups. Statistics were calculated using unpaired Wilcoxon test.

Finally, we assessed whether the differential vaccine responses of CD4 and CD8 T cell subsets noted when splitting the MS-aCD20 patients by RBD IgG response were related to the induction of antigen-specific T cell responses. The MS-aCD20 RBD Ab-group showed markedly lower abundance of AIM^+^ CD4 T cells and especially AIM^+^ cTfh cells at timepoints T4 and T5 (Figure 6F). In contrast, AIM^+^ Th1 cells were similar in RBD Ab+ and RBD Ab-groups. Notably, AIM^+^ CD8 T cell vaccine responses were significantly more robust in MS-aCD20 RBD Ab-patients compared to RBD Ab+ patients after the second vaccine dose, supporting the notion that SARS-CoV-2 mRNA vaccine-induced CD8 T cell responses are more vigorous in patients who lack B cells and a productive antibody response due to aCD20 treatment. Thus, these analyses indicate that the subset of anti-CD20 treated MS patients who failed to mount anti-RBD IgG responses exhibited more substantial perturbations in their vaccine-induced cellular immune response. Together, these data underscore the interrelated and coordinated nature of mRNA vaccine induced immune responses, provide insights into generation of T cell responses even in the absence of humoral immunity, and shed light on underlying “immune health” differences in MS patients on aCD20 therapy.

## Discussion

Immunocompromised patients and patients with systemic autoimmunity were not included in the two phase-3 clinical trials that assessed the BNT162b2 (Pfizer-BioNTech) and mRNA-1273 (Moderna) SARS-CoV-2 mRNA vaccines^4, 5^. There is currently a pressing need to understand immune responses induced by mRNA vaccines in these populations, since such insights could inform clinical practice guidelines and mitigation measures as the healthy population is gradually returning to pre-pandemic norms of social interaction. Moreover, there is relatively little information available on the immune mechanisms of mRNA vaccine-induced immune responses in settings of immune perturbation or immune suppression. Insights gained from such analyses may help improve strategies aimed at achieving protective immunological memory in vulnerable populations. Our goal in this study was to evaluate the impact of aCD20 therapy on SARS-CoV-2 mRNA vaccine responses. Therapy with aCD20 is used in many clinical settings including cancer immunotherapy, rheumatology and neurology. In MS, aCD20 treatment is commonly used as monotherapy. This offers the advantage that studying this patient population is relatively less confounded by other concurrent immune therapies.

Examining vaccine-induced immune responses in MS-aCD20 patients led to several key observations. First, MS patients on aCD20 therapy had reduced B cell functional responses that included limited antibody induction to Spike and RBD, as well as poor generation of antigen-specific memory B cells. Some MS-aCD20 patients did generate antibody to Spike and RBD and these responses correlated with detection of B cells in the blood. There were, however, some patients who made antibody responses to Spike in the absence of detectable circulating B cells, perhaps pointing to early repopulation of B cells within the lymphoid tissue that are capable of contributing to serological responses. Thus, although antibody responses were compromised in patients on aCD20 therapy, some antibody generation occured in a subset of patients, perhaps related to time since last treatment with aCD20.

SARS-CoV-2 mRNA vaccines also induce T cell responses^44, 70, 71^. Although induction of neutralizing antibodies is likely to be important in vaccine-induced protection, precise correlates of immunity remain to be completely defined and recent evidence also points to a role of T cells in mitigating severe disease upon infection^40–43, 72^. Despite poor antibody responses in most MS-aCD20 patients, all of these patients generated robust CD4 and CD8 T cell responses to SARS-CoV-2 mRNA vaccination suggesting that vaccinating such subjects on B cell immunosuppression is likely to provide some measure of immunity to SARS-CoV-2. Despite this robust T cell priming, MS-aCD20 patients had selective defects in maintaining similar frequencies of antigen-specific cTfh cells compared to HCs. The defects in cTfh responses were even more dramatic in MS-aCD20 who did not generate RBD antibody responses. While it is possible that some of these changes could reflect an impact of aCD20 on a subset of CD20^+^ T cells^73, 74^, these data are also consistent with the idea that not only do Tfh cells provide help to B cells^75^, but that germinal center B cells also augment, or are necessary for, optimal and sustained Tfh cell responses^76^. In contrast to the cTfh responses, Th1 cell priming and maintenance were only midly impacted and CD8 T-cell responses were augmented, especially following the second vaccine dose. Thus, although the clinical correlates of protection from SARS-CoV-2 infection remain incompletely understood, MS-aCD20 patients appear able to mount productive antiviral CD8 and Th1 CD4 responses, despite suboptimal (or even undetectable) antibody responses. Recent studies in cancer patients indicate an association between T cell responses and better resolution of COVID-19^43^. Thus, it is possible that these vaccine induced T cell responses may confer some protection from severe COVID-19 outcomes in these aCD20-treated patients. In the absence of robust antibody responses, however, these patients may not achieve as rapid sterilization from SARS-CoV-2 infection, potentially resulting in longer carrier states and/or increased risk of transmission.

B cell reconstitution in the circulation was, as expected, preferentially detected in patients who were farther removed from their last aCD20 treatment. This patient subgroup was more successful at generating both antibodies and memory B cells against Spike and RBD. In addition, these patients had less perturbed CD4 and CD8 T cell response profiles to mRNA vaccination. The magnitude of vaccine-induced humoral responses correlated better with the extent of B cell reconstitution at the time of vaccination than with the time window between the vaccination and the last aCD20 infusion, arguing that the underlying mechanism for this effect is B cell reconstitution. Although some MS-aCD20 patients made antibodies to Spike in the absence of detectable circulating B cells, stronger antibody responses to Spike and RBD were associated with detectable B cells in the blood. Thus, assessing re-emergence of peripheral B cells may be a better marker than time since last aCD20 treatment to determine which patients will generate humoral immunity following vaccination. While requiring confirmation in larger cohorts, our results provide insights that may contribute to the development of clinical practice guidelines, including considerations around the most appropriate timing for administering SARS-CoV-2 vaccines or boosters in patients on aCD20 therapies.

One unexpected finding was the more robust vaccine-induced CD8 T cell response after SARS-CoV-2 mRNA vaccination in the MS-aCD20 group, especially the patients who failed to generate anti-RBD IgG. This difference was most prominent following the administration of the second vaccine dose. On one hand, these findings are evidence of effective immune priming by mRNA vaccines in the absence of circulating B cells, an observation with obvious implications for protection from severe COVID-19 in vulnerable populations including cancer and rheumatology patients vaccinated against SARS-CoV-2. However, these observations may also be relevant for application of mRNA vaccines in other settings such as neoantigen cancer vaccines in patients with B cell deficiencies^77^. An important question to address in the future is the underlying mechanism of this augmented CD8 T cell response. One possibility is that, in the absence of antibody, there is an increased abundance of antigen to drive CD8 T cell activation and proliferation due to lack of clearance by vaccine-induced antibodies. Alternatively, B cells may play a direct role in attenuating CD8 T cell responses^78, 79^. For example, regulatory B cells, possibly primed by the initial dose of mRNA vaccine, may limit CD8 T cell responses following the second dose. In this scenario, distinguishing the role of B cells in fostering effective priming of CD4 T cells from negatively regulating CD8 T cells may be important for optimizing future mRNA vaccination approaches in other settings. A third possibility is through effects of antibody or immune complexes via engagement of the inhibitory Fc receptor FcγRIIB on dendritic cells^80, 81^ or CD8 T cells^82^. In the absence of Spike-containing immune complexes in MS-aCD20 patients, CD8 T cell responses may be augmented specifically after the second dose when such FcγRIIB-mediated inhibition could occur in the HC cohort. Future studies will be necessary to determine the contribution of these possible mechanisms. Nevertheless, the augmented CD8 T cell response in MS-aCD20 documented here may have implications for future protective immunity and provide impetus to further interrogate CD8 T cell responses in other immunocompromised populations.

Overall, these studies provide strong evidence of immune priming by SARS-CoV-2 mRNA vaccines in MS-aCD20 patients. Although most of these patients do not generate optimal antibody responses, T cell priming, especially of Th1 and CD8 T cells, remains largely intact. Treatment with aCD20 and B cell deficiency were associated with altered coordination of the immune response, however, and cTfh responses were compromised. Nevertheless, despite the intent of aCD20 treatment to remove B cell mediated immunity in patients with MS, including effects of B cells in presenting antigen to CD4 T cells, these studies reveal variable levels of residual underlying immune functionality in MS-aCD20 patients. The analysis of mRNA vaccine induced immune responses not only to measure immunity to SARS-CoV-2 but also as an “analytical vaccine” offers insights into the underlying immune health and fitness of MS-aCD20 patients. Overall, these data provide key insights about the ability to generate immune responses in immunocompromised populations that will be relevant for clinical guidance in these patients and possible public health recommendations for vulnerable populations.

## Methods

### Study design

In this longitudinal study, healthy controls (HC, n=10) and MS patients treated with anti-CD20 (MS-αCD20, n=20, 19 patients on ocrelizumab and 1 patient on rituximab) were recruited between December 2020 and April 2021. Plasma and peripheral blood mononuclear cells (PBMC) were collected immediately prior to the 1^st^ vaccine dose (T1), 10∼12 days post 1^st^ vaccine dose (T2), immediately prior to the 2^nd^ vaccine dose (T3), 10∼12 days post 2^nd^ vaccine dose (T4) and 25∼30 days following the 2^nd^ vaccine dose (T5). Clinical information for HC and MS-aCD20 vaccinees can be found in **Extended Data Table 1**. All experiments were conducted in a blinded fashion with designated members of the clinical team (that were not part of running the assays) having access to the sample key until the data were collected, at which point all researchers were unblinded for the analysis. All subjects enrolled in this study provided informed consent as part of protocols approved by the University of Pennsylvania Institutional Review Boards (IRBs) and in compliance with the Declaration of Helsinki principles.

### Cell isolation and cryopreservation

Venous blood was collected in multiple 10ml K2 EDTA tubes (BD Vacutainer, Cat # 366643). Blood was diluted at 1:1 ratio with PBS that contains 2mM EDTA and then slowly transferred to a 50ml tube that contained 15ml Ficol (GE Healthcare, Cat # CA95038-168L). Tubes were then spun at 700g at room temperature with no brake. PBMC layers were collected using a transfer pipet and then washed once with 40ml PBS+EDTA buffer before submitted for cell count. Cells were then resuspended in freezer media (Human AB serum+10% DMSO) and aliquoted into cryopreserved tubes (∼20million per tube). PBMC samples were first stored in Mr. Frosty freezing containers at −80°C and then transferred to liquid nitrogen tanks for long term storage.

### Plasma isolation

Venous blood was collected in a 10ml K2 EDTA tube (BD Vacutainer, Cat # 366643). The tube was then stored upright at room temperature for 30min before centrifugation at 4°C for 10 at 2500g (with swinging bucket rotor). Supernatants were then collected, aliquoted and stored at −80°C until further use.

### Detection of SARS-CoV-2-Specific Antibodies

Plasma samples were tested for SARS-CoV-2-specific antibody by enzyme-linked immunosorbent assay (ELISA) as previously described^83^. The estimated sensitivity of the test is 100% [95% confidence interval (CI), 89.1 to 100.0%], and the specificity is 98.9% (95% CI, 98.0 to 99.5%). Plasmids encoding the recombinant full-length spike protein and the RBD were provided by F. Krammer (Mt. Sinai) and purified by nickel-nitrilotriacetic acid resin (Qiagen). Monoclonal antibody CR3022 was included on each plate to convert OD values into relative antibody concentrations. Plasmids to express CR3022 were provided by I. Wilson (Scripps).

### Flow Cytometry

Samples were acquired on a 5 laser BD FACS Symphony A5 (X50 SORP). Standardized SPHERO rainbow beads (Spherotech, Cat#RFP-30-5A) were used to track and adjust PMTs over time. UltraComp eBeads (ThermoFisher, Cat#01-2222-42) were used for compensation. Up to 1×10^6^ PBMCs were acquired per each sample. All antibodies used for high-dimensional FACS analysis can be found in **Extended Data Table 4**.

### Detection of SARS-CoV-2 specific Memory B Cells

Antigen-specific B cells were detected using biotinylated proteins in combination with different streptavidin (SA)-fluorophore conjugates, as previously described^51^. Briefly, biotinylated proteins were multimerized with fluorescently labeled SA for 1 hour at 4C. Full-length spike protein (R&D Systems) was mixed with SA-BV421 (Biolegend) at a 10:1 mass ratio (e.g., 200ng spike with 20ng SA; ∼4:1 molar ratio). Spike RBD (R&D Systems) was mixed with SA-APC (Biolegend) at a 2:1 mass ratio (e.g., 25ng RBD with 12.5ng SA; ∼4:1 molar ratio). Biotinylated influenza HA pools (A/Brisbane/02/2018/H1N1, B/Colorado/06/2017; Immune Technology) were mixed with SA-PE (Biolegend) at a 6.25:1 mass ratio (e.g., 100ng HA pool with 16ng SA; ∼6:1 molar ratio). Excess biotin was subsequently removed using Zebra Spin Desalting Columns 7K MWCO (Thermo Fisher) and protein was quantified with a Pierce BCA Assay (Thermo Fisher). SA-BV711 (BD Bioscience) was used as a decoy probe without biotinylated protein to gate out cells that non-specifically bind streptavidin. Antigen probes for spike, RBD, and HA were prepared individually and mixed together after multimerization with 5uM free D-biotin (Avidity LLC) to minimize potential cross-reactivity between probes.

### Activation induced marker (AIM) assays

PBMCs were thawed by diluting with 10mL of warm RPMI supplemented with 10% FBS, 2mM L-Glutamine, 100 U/mL Penicillin, and 100 mg/mL Streptomycin (R10) and washed once in R10. Cell counts were obtained with a Countess automated cell counter (Thermo Fisher), and each sample was resuspended in fresh R10 to a density of 5×10^6^ cells/mL. For each condition, duplicate wells containing 1×10^6^ cells in 200mL were plated in 96-well round-bottom plates and rested overnight in a humidifed incubator at 37°C, 5% CO_2_. After the rest, CD40 blocking antibody (0.5mg/mL final concentration) was added to cultures for 15 minutes prior to stimulation, and cells were subsequently stimulated for 24 hours with costimulation (anti-human CD28/CD49d, BD Biosciences) and peptide megapools (CD4-S for all CD4 T cell analyses, CD8-E for all CD8 T cell analyses) at a final concentration of 1 mg/mL. Peptide megapools were prepared as previously described^66, 67^. Matched unstimulated samples for each donor at each timepoint were treated with costimulation alone. 20 hours post-stimulation, antibodies targeting CXCR3, CCR7, CD40L, CXCR5, and CCR6 were added to the culture along with monensin (GolgiStop, BD Biosciences) for a four hour stain at 37°C. After four hours, duplicate wells were pooled and cells were washed in PBS supplemented with 2% FBS (FACS buffer). Cells were stained for 10 minutes at room temperature with Ghost Dye Violet 510 and Fc receptor blocking solution (Human TruStain FcX™, BioLegend) and washed once in FACS buffer. Surface staining for 30 minutes at room temperature was then performed with antibodies directed against CD4, CD8, CD45RA, CD27, CD3, CD40L, CD200, and 41BB in FACS buffer. Cells were washed once in FACS buffer, fixed and permeabilizied for 30 minutes at room temperature (eBioscience™ Foxp3 / Transcription Factor Fixation/Permeabilization Concentrate and Diluent), and washed once in 1X Permeabilization Buffer prior to staining for intracellular IFN-γ overnight at 4°C. Cells were then washed once and resuspended in 1% paraformaldehyde in PBS prior to data acquisition.

All data from AIM expression assays were background-subtracted using paired unstimulated control samples. For memory T cell and helper T cell subsets, the AIM+ background frequency of non-naïve T cells was subtracted independently for each subset. AIM^+^ cells were identified from non-naïve T cell populations. AIM^+^ CD4 T cells were defined by dual-expression of CD200 and CD40L. AIM^+^ CD8 T cells were defined by dual-expression of 41BB and intracellular IFN-γ.

### High-dimensional data analysis of flow cytometry data

Opt-SNE and FlowSOM analyses were performed using OMIQ (https://app.omiq.ai/). Total CD4, activated CD4 and activated CD8 T cells were analyzed separately. Markers used for all three analyses: CD27, CD45RA, CD127, T-bet, CXCR5, CD71, CD38, CCR6, HLA-DR, CTLA-4, PD-1, CCR7, CD25, CXCR3, ICOS, CXCR4, Foxp3, Ki67.

The Opt-SNE parameters were:

- Total CD4 T cells: max iterations 1000, perplexity 30, theta 0.5, seed 1234. Subsampling equal between cohorts: total 3M cells (1.5M for HC and 1.5M for MS-aCD20 groups)
- Activated Ki67^+^CD38^+^ CD4 T cells: max iterations 1000, perplexity 30, theta 0.5, components 2, seed 1234. Subsampling equal between cohorts: total 13822 cells (6911 cells for HC and 6911 cells for MS-aCD20 groups)
- Activated Ki67^+^CD38^+^ CD8 T cells: max iterations 1000, perplexity 30, theta 0.5, components 2, seed 1234. Subsampling equal between cohorts: total 54446 cells (27223 cells for HC and 27223 cells for MS-aCD20 groups)

FlowSOM was performed in all three analyses using the same markers outlined above for opt-SNE and with the following parameters: number of clusters 100, number of metaclusters 10 (activated CD4, activated CD8 T cells) or 15 (total CD4 T cells), distance metric Euclidean and consensus metaclustering.

To group individual samples on the basis of their T cell landscape, pairwise EMD values were calculated on the opt-SNE axes for all HC and MS-aCD20 vaccinees at all timepoints collected using the *emdist* package in R, as previously described^68, 69^.

### Statistical analysis

Owing to the heterogeneity of clinical and flow cytometric data, nonparametric tests of association were preferentially used throughout this study unless otherwise specified. Correlation coefficients between ordered features (including discrete ordinal, continuous scale, or a mixture of the two) were quantified by the Spearman rank correlation coefficient, and significance was assessed by the corresponding nonparametric methods (null hypothesis: ρ = 0). Tests of association between mixed continuous versus nonordered categorical variables were performed by unpaired Wilcoxon test (for *n* = 2 categories) or Kruskal-Wallis test (for *n* > 2 categories). Association between categorical variables was assessed by Fisher’s exact test. All tests were performed in a two-sided manner, using a nominal significance threshold of *P* < 0.05 unless otherwise specified. Other details, if any, for each experiment are provided within the relevant figure legends. For *p* values: * indicates *p* < 0.05, ** indicates *p* < 0.01, *** indicates *p* < 0.001, **** indicates *p* < 0.0001.

## Data Availability

All data are available in the main text or the supplementary materials. Raw data files and reagents are available from the authors upon request.

## Acknowledgments

We would like to thank the study participants for their generosity in making the study possible. We also thank the members of the Wherry and Bar-Or labs for helpful discussions and feedback. This work was supported by grants from the NIH AI105343, AI082630, AI108545, AI155577, AI149680 (to EJW), AI152236 (to PB), P30-AI0450080 (to ETLP), R01 AI118694 and UC4 DK112217 (to M.R.B.), T32 AR076951-01 (to SAA), T32 CA009140 (to DM), U19AI082630 (to SEH and EJW), UM1 AI144288 (ABO), NMSS SI-2011-37160 (ABO), funding from the Allen Institute for Immunology (to SAA, EJW), Chen Family Research Fund (to SAA), the National Multiple Sclerosis Society-American Brain Foundation Clinician Scientist Award (to MK), the Parker Institute for Cancer Immunotherapy (to EJW), the Penn Center for Research on Coronavirus and Other Emerging Pathogens (to PB), the University of Pennsylvania Perelman School of Medicine COVID Fund (to RRG, EJW), the University of Pennsylvania Institute for Immunology Glick COVID-19 research award (to M.R.B.), the University of Pennsylvania Perelman School of Medicine 21^st^ Century Scholar Fund (to RRG), and a philanthropic gift from Jeffrey Lurie, Joel Embiid, Josh Harris, and David Blitzer (to SEH). Work in the Wherry lab is supported by the Parker Institute for Cancer Immunotherapy. This work was also supported by NIH contract Nr. 75N9301900065 (to DW, AS).

## Author contributions

EJW, RL and ABO concieved the study. SAA, MMP, RRG, RL, DM, SG and KAL carried out experiments. MK, DJ, CM, RL and ABO were invovled in clinical recruitment. RL, AR and JK processed peripheral blood samples and managed the sample database. DW, AG and AS provided data and materials. ARG, EJW, RL and ABO supervised the study. All authors participated in data analysis, interpretation and manuscipt review. SAA, EJW, RL and ABO wrote the manuscript.

## Extended Data Figure legends

**Extended Data Figure 1. A-B)** Spearman correlation analysis between the weeks elapsed since last infusion administration with anti-Spike IgG **(A)** or anti-RBD IgG **(B)** at T5 for MS-aCD20 patients. **C)** Gating strategy and representative plots for flow cytometric analysis of total B cells. **D)** Gating strategy and representative plots for flow cytometric analysis of SARS-CoV-2-specific memory B cells. Cells were stained with fluorescently labeled SARS-CoV-2 full-length spike protein, SARS-CoV-2 spike receptor binding domain (RBD), and influenza hemagglutinin (HA). Spike^+^ HA^-^ cells were subsequently analyzed for binding to RBD.

**Extended Data Figure 2.** Gating strategy for FACS-based analysis of CD4 and CD8 T cells following SARS-CoV-2 mRNA vaccination

**Extended Data Figure 3. A)** Opt-SNE projections of concatenated cytometry data for total CD3^+^CD4^+^ T cells for each timepoint and group combination are shown. **B)** Surface expression intensity of the markers indicated on the opt-SNE 2D-map generated with all samples in A. **C)** FlowSOM metaclusters were created using total CD3^+^CD4^+^ T cells concatenated from all samples and projected to the opt-SNE map. **D)** Surface expression intensity heatmap of the markers indicated for each of the 15 FlowSOM metaclusters in C. **E)** Volcano plots of the logFC (log fold change) of the abundance between the HC and MS-aCD20 groups for each of the 15 metaclusters indicated in C and the −log10 value of the false discovery rate (FDR) for timepoints 2 and 4.

**Extended Data Figure 4. A)** The abundance of metaclusters 2, 4, 5, 6, 8, 9 and 10 as percentage of activated Ki67^+^CD38^+^ CD4 T cells is shown for HC (grey) and MS-aCD20 (orange) groups; unpaired Wilcoxon test *p* values are shown when *p* < 0.05 between groups. **B)** The abundance of metacluster 3 as percentage of total non-naive CD4 T cells is shown for HC (grey) and MS-aCD20 (orange) groups; unpaired Wilcoxon test *p* values are shown when *p* < 0.05 between groups.

**Extended Data Figure 5. A)** Gating strategy for identifying T cell subsets. **B)** Timepoint 4 AIM^+^ CD4 T cell (left) and CD8 T cell (right) frequency in whole PBMCs or PBMCs depleted of B cells by magnetic separation. Values represent the background-subtracted frequency of AIM^+^ non-naïve CD4 T cells above paired baseline frequencies. Lines connect paired samples from individual donors. Gray indicates HC; orange indicates patients with MS receiving anti-CD20 therapy. **C)** Frequency of memory CD4 T cell subsets in total non-naïve CD4. Left panels depict the percent of total non-naïve T cells that are in each subset. Right panels depict the relative frequency of each memory T cell subset in the total non-naive population. CM = CD45RA^-^CD27^+^CCR7^+^, EM1 = CD45RA^-^CD27^+^CCR7^-^, EM2 = CD45RA^-^CD27^-^CCR7^+^, EM3 = CD45RA^-^CD27^-^CCR7^-^, EMRA = CD45RA^+^CD27^-^CCR7^-^. **D)** Frequency of T helper subsets in total non-naïve CD4 T cells. Left panel depicts the percent of total non-naïve T cells that are helper T cells in each subset. Right panel depicts the relative frequency of each helper T cell subset in the total non-naive population. cTfh = CXCR5^+^, Th1 = CXCR5^-^CXCR3^+^CCR6^-^, Th2 = CXCR5^-^CXCR3^-^CCR6^-^, Th17 = CXCR5^-^CXCR3^-^CCR6^+^, Th1/17 = CXCR5^-^ CXCR3^+^CCR6^+^. **E**) Summary data of AIM+ frequencies of the indicated T cell populations following vaccination. Values represent the background-subtracted frequency of AIM^+^ non-naïve T cells above paired baseline frequencies. Statistics were calculated using unpaired Wilcoxon test. Gray indicates healthy controls; orange indicates subjects with MS receiving anti-CD20 therapy.

**Extended Data Figure 6. A)** The abundance of metaclusters 1, 2, 3, 4, 5, 9 and 10 as percentage of activated Ki67^+^CD38^+^ CD8 T cells is shown for HC (grey) and MS-aCD20 (orange) groups; unpaired Wilcoxon test *p* values are shown when *p* < 0.05 between groups.

**Extended Data Figure 7.** Frequency of memory CD8 T cell subsets in total non-naïve CD8 T cells. Left panels depict the percent of total non-naïve T cells that are in each subset. Right panels depict the relative frequency of each memory T cell subset in the total non-naive population. CM = CD45RA^-^CD27^+^CCR7^+^, EM1 = CD45RA^-^CD27^+^CCR7^-^, EM2 = CD45RA^-^CD27^-^CCR7^+^, EM3 = CD45RA^-^CD27^-^CCR7^-^, EMRA = CD45RA^+^CD27^-^CCR7^-^.

**Extended Data Figure 8. A)** Boxplots representing the summary statistics (median, distribution) of the EMD distances to MS-aCD20 RBD IgG-samples on the activated CD4 T cell opt-SNE maps across all timepoints T1-T5 for the three groups: Healthy RBD Ab+, MS-aCD20 RBD Ab+ and MS-aCD20 RBD Ab-. Pairwise comparisons of means were done with Wilcoxon test and *p* values are shown. **B)** Boxplots representing the summary statistics (median, distribution) of the EMD distances to Healthy RBD Ab+ samples on the activated CD8 T cell opt-SNE maps across all timepoints T1-T5 for the three groups: Healthy RBD IgG+, MS-aCD20 RBD Ab+ and MS-aCD20 RBD Ab-. Pairwise comparisons of means were done with Wilcoxon test and *p* values are shown.

