## Supplementary_materials for "Altered cellular and humoral immune responses following SARS-CoV-2 mRNA vaccination in patients with multiple sclerosis on anti-CD20 therapy"

**Supplementary Table 1.**

| | MS (n=20) | HC (n=10) | *p* - value |
|---|---|---|---|
| **Age**, mean years± SD (range) | 40.35±8.44 [27-57] | 35.2±9.8 [25-61] | p=0.146 |
| **Females,** n (%) | 15 (75) | 6 (60) | p=0.431 |
| **Relapsing Remitting MS,** n (%) | 21(100) | - | |
| **Vaccine type** | | | |
| Pfizer, n (%) | 12 (61.9) | 8 (80) | p=0.419 |
| Moderna, n (%) | 8 (38.1) | 2 (20) | |
| **Time from last aCD20 to first vaccine (weeks),** mean±SD [range] | 19.77±9.52 [2.6-41.1] | - | |
| **Prior cycles of aCD20**, mean±SD [range] | 3.2±1.6 [1-7] | - | |
| **Type of aCD20** | | | |
| Ocrelizumab, n (%) | 19 (95) | - | |
| Rituximab, n (%) | 1 (5) | - | |
| **EDSS**, mean±SD [range] | 1.5±2.2 (0-6.5) | - | |

**Supplementary Table 1.** MS-aCD20 and HC cohort Demographics.

**Supplementary Table 2.**

| | HC | MS-αCD20 | *p* - value |
|---|---|---|---|
| **anti-Spike IgG** | | | |
| T2 | 100% | 28.57% | 0.0006 |
| T3 | 100% | 29.41% | 0.0004 |
| T4 | 100% | 82.35% | 0.2735 |
| T5 | 100% | 88.89% | 0.5238 |
| **RBD Protein IgG** | | | |
| T2 | 60% | 7.14% | 0.0088 |
| T3 | 100% | 17.65% | <0.0001 |
| T4 | 100% | 41.48% | 0.0031 |
| T5 | 100% | 50% | 0.0098 |

**Supplementary Table 2.** Anti-Spike and anti-RBD IgG serological positivity rates in HC and MS-aCD20 patients across different time points. Two-sided Fisher's exact test.

**Supplementary Table 3.**

| Antigen-specific memory B cells | Timepoint | Comparison | p - value | method |
|---|---|---|---|---|
| $Spike^{+}$ | T2 | HC vs MS-aCD20 | 9.20E-06 | Wilcoxon |
| $Spike^{+}$ | T3 | HC vs MS-aCD20 | 1.10E-05 | Wilcoxon |
| $Spike^{+}$ | T4 | HC vs MS-aCD20 | 2.60E-05 | Wilcoxon |
| $Spike^{+}$ | T5 | HC vs MS-aCD20 | 1.10E-05 | Wilcoxon |
| $RBD^{+}$ | T2 | HC vs MS-aCD20 | 5.50E-06 | Wilcoxon |
| $RBD^{+}$ | T3 | HC vs MS-aCD20 | 3.30E-06 | Wilcoxon |
| $RBD^{+}$ | T4 | HC vs MS-aCD20 | 1.50E-05 | Wilcoxon |
| $RBD^{+}$ | T5 | HC vs MS-aCD20 | 8.70E-06 | Wilcoxon |

**Supplementary Table 3.** The frequencies of Spike+ and RBD+ antigen-specific memory B cells were compared between HC and MS-aCD20 groups at each timepoint. Wilcoxon signed-rank testing was used, and the *p* values are shown.

**Supplementary Table 4.**

| Antibody reactivity | Fluorochrome Conjugation | Clone | Supplier | Catalog # |
|---|---|---|---|---|
| CD27 | BUV 395 | L128 | BD | 563815 |
| CD71 | BUV 496 | M-A712 | BD | 750652 |
| CD3 | BUV 563 | UCHT1 | BD | 748569 |
| CD8 | BUV 615 | RPA-T8 | BD | 751518 |
| CD38 | BUV 661 | HIT2 | BD | 612969 |
| CCR6 | BUV 737 | 11A9 | BD | 612780 |
| HLA-DR | BUV 805 | G46-6 | BD | 748338 |
| CTLA4 | BV 421 | BNI3 | BD | 562743 |
| PD-1 | BV 480 | EH12.11 | BD | 566112 |
| CCR7 | BV 510 | G043H7 | Biolegend | 353232 |
| Zombie Yellow | BV 570 | - | Biolegend | 423103 |
| CD45RA | BV 605 | HI100 | Biolegend | 304134 |
| CD25 | BV 650 | M-A251 | BD | 563719 |
| CXCR3 | BV 711 | G025H7 | Biolegend | 353732 |
| CD4 | BV 750 | SK3 | BD | 566355 |
| ICOS | BV 785 | C398.4A | Biolegend | 313534 |
| SLAM | AF 488 | A12 (7D4) | Biolegend | 306312 |
| CD127 | BB 700 | HIL-7R-M21 | BD | 566398 |
| CXCR4 | PE-Cy5 | 12G5 | Biolegend | 306508 |
| FoxP3 | PE-Cy5.5 | PCH101 | Fisher | 35-4776-42 |
| Ki67 | PE-Cy7 | B56 | BD | 561283 |
| T-bet | AF 647 | 4B10 | Biolegend | 644804 |
| CXCR5 | APC-R700 | RF8B2 | BD | 565191 |
| Bcl-6 | APC-Cy7 | K112-91 | BD | 563581 |

**Supplementary Table 4.** Antibody specifications used for FACS-based high-dimensional analysis.

### Extended Data Figure 1.

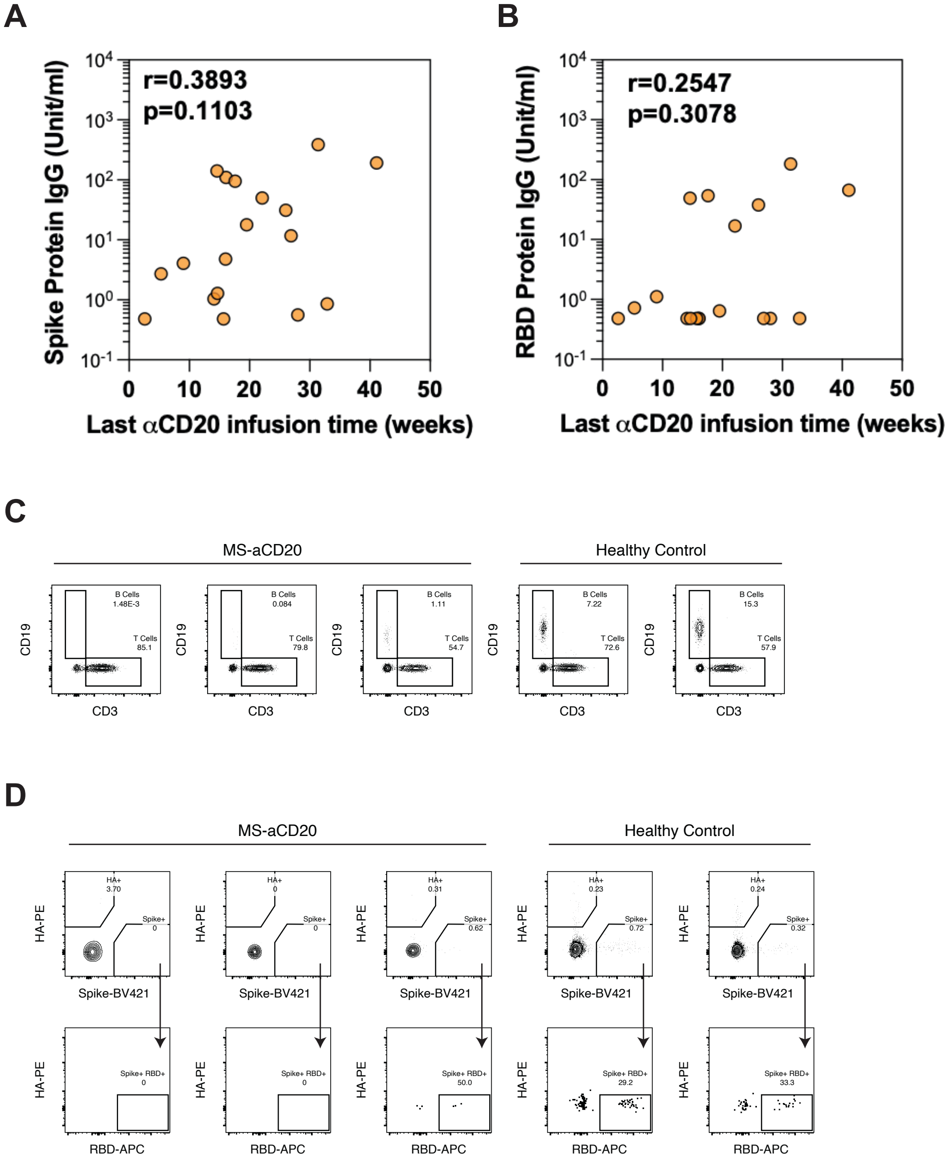

### Extended Data Figure 2.

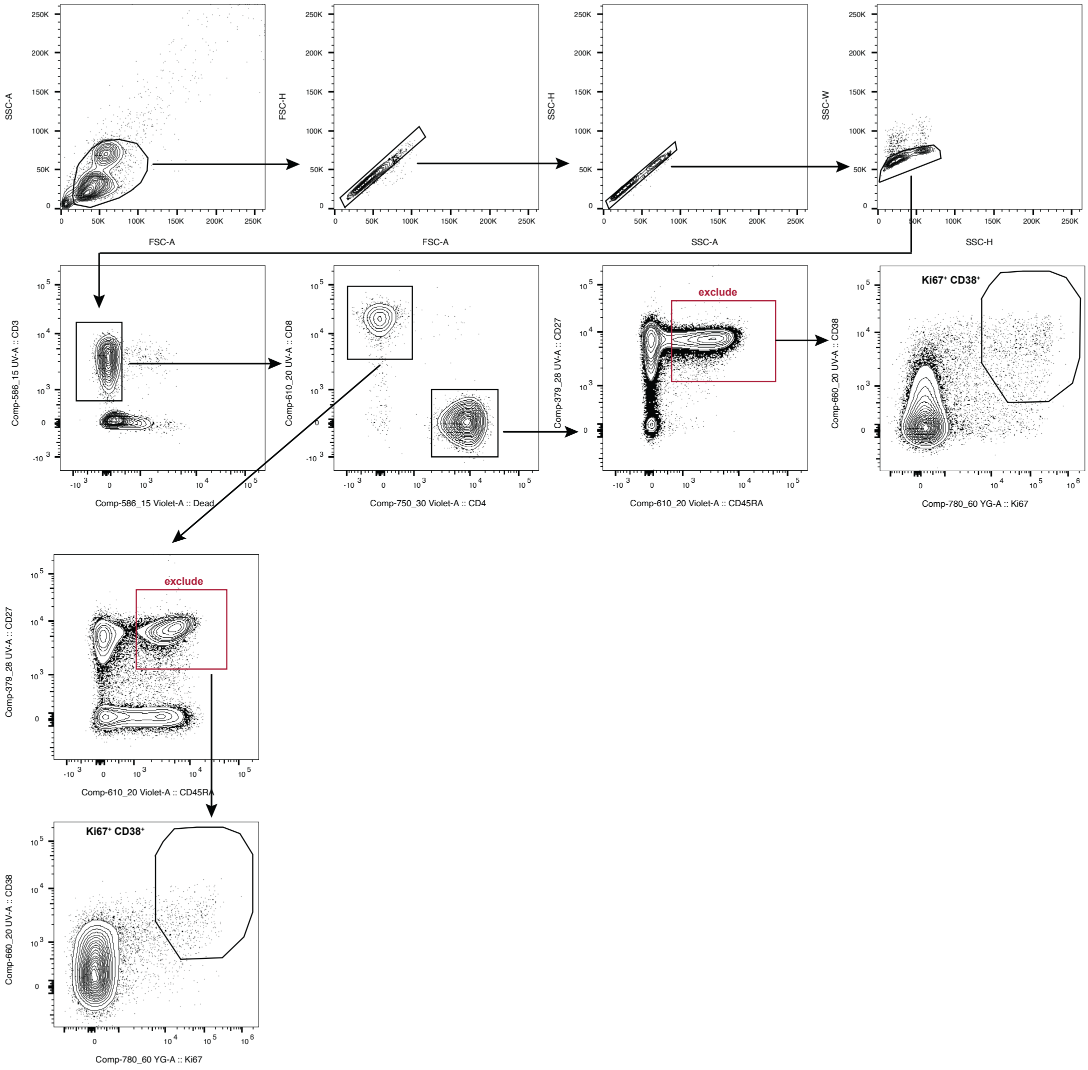

### Extended Data Figure 3.

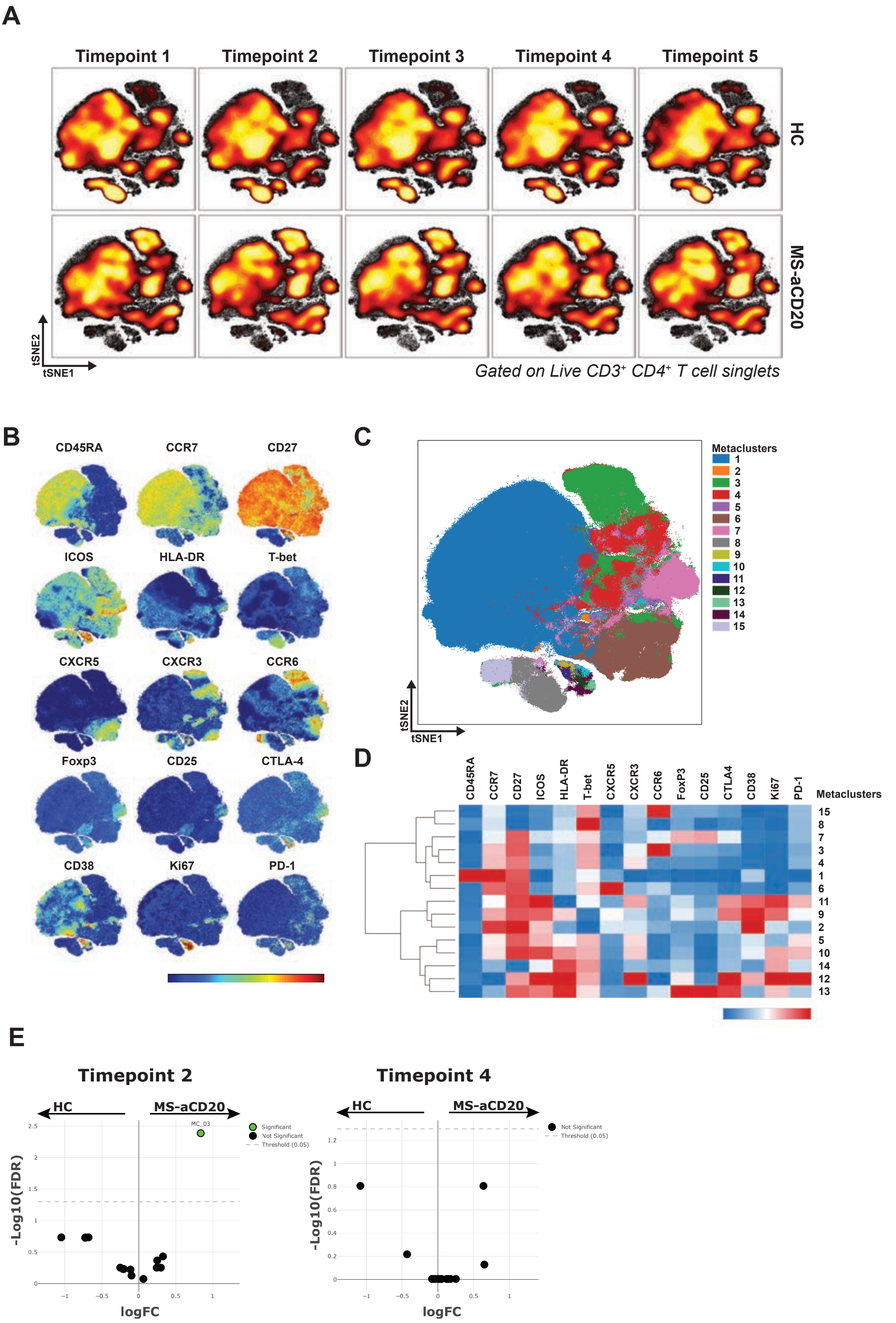

**Extended Data Figure 4.**

**A**

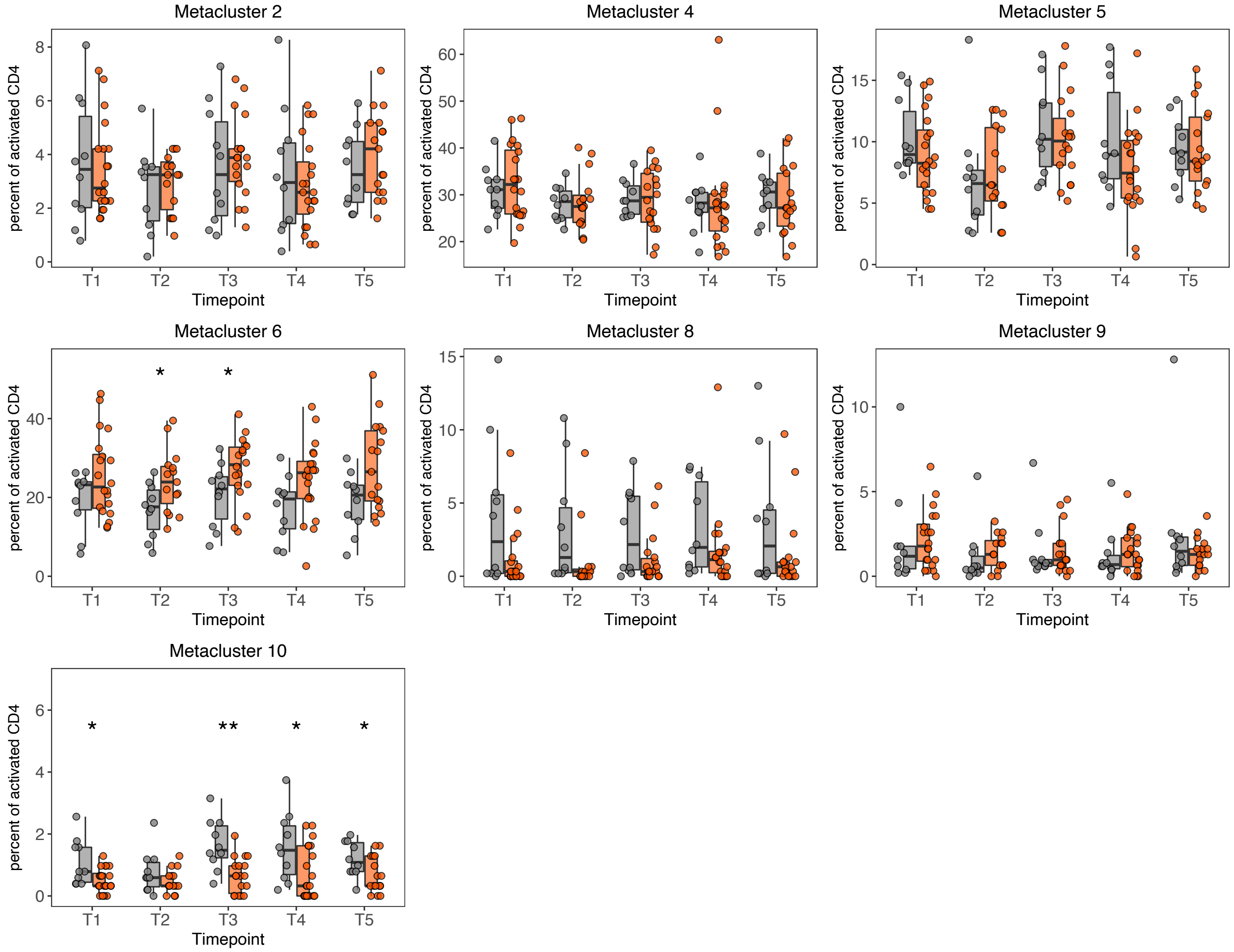


**B**

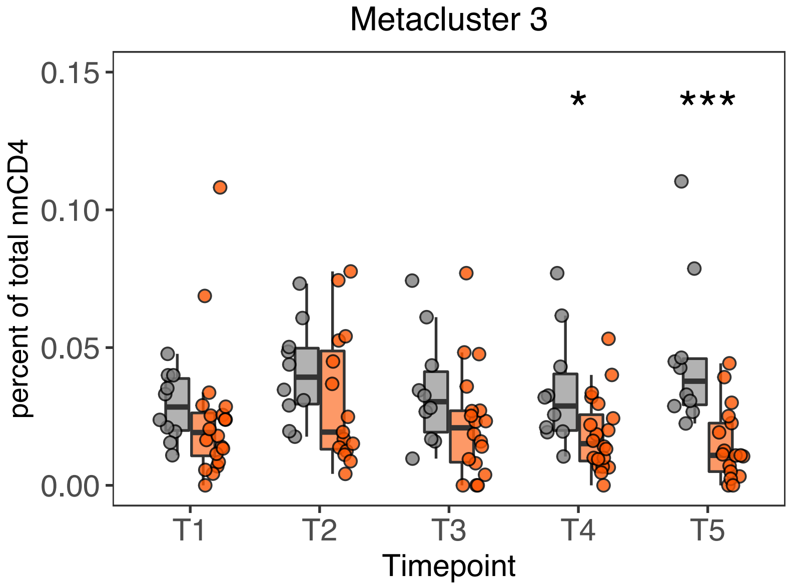

#### Extended Data Figure 5.

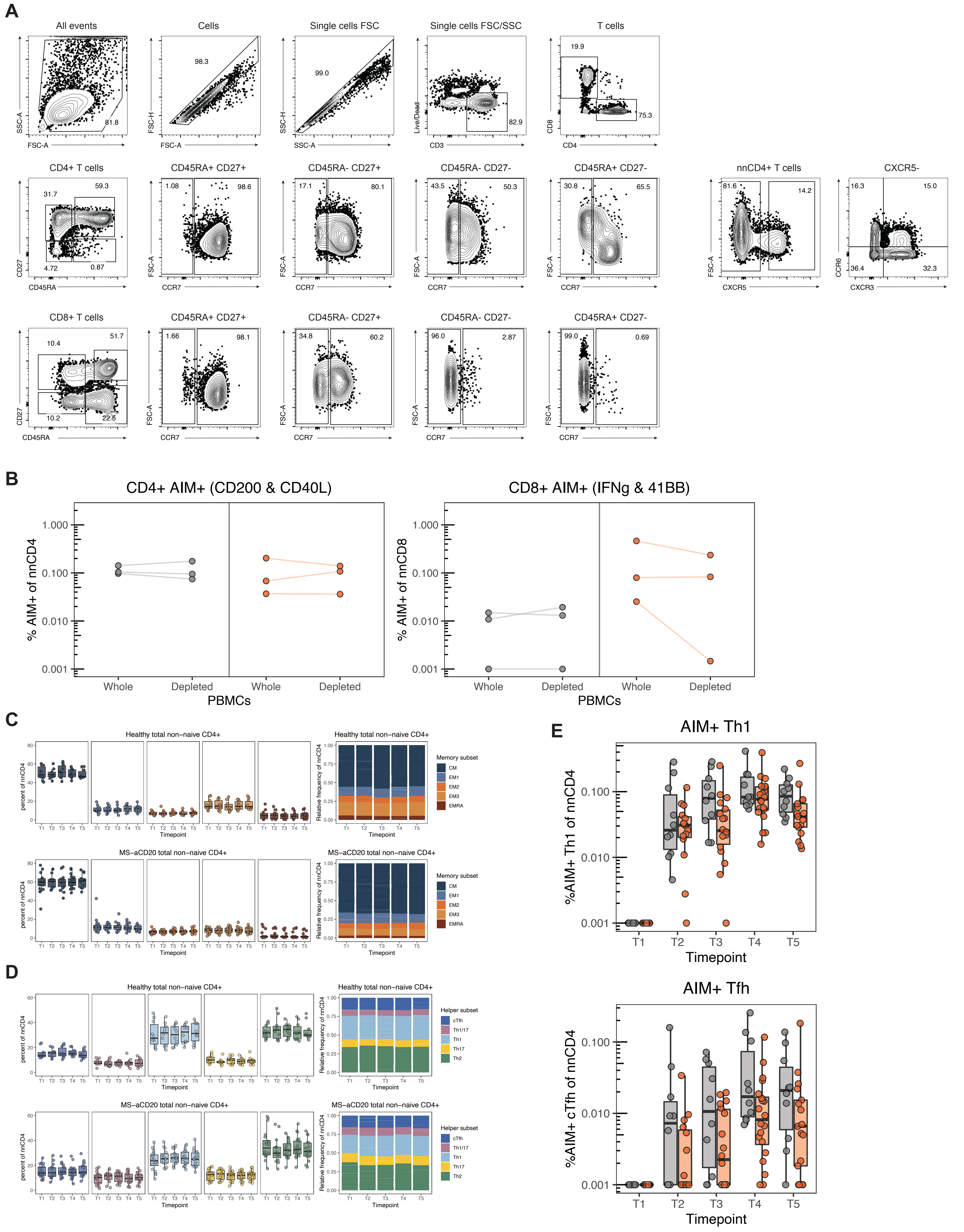

### Extended Data Figure 6.

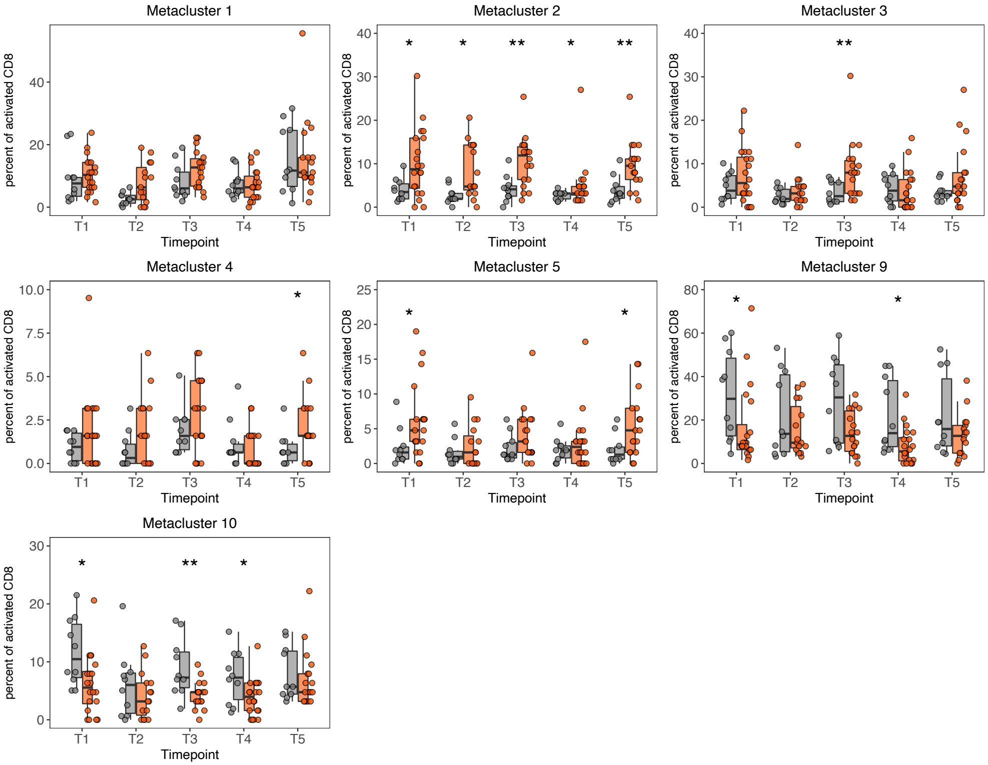

### Extended Data Figure 7.

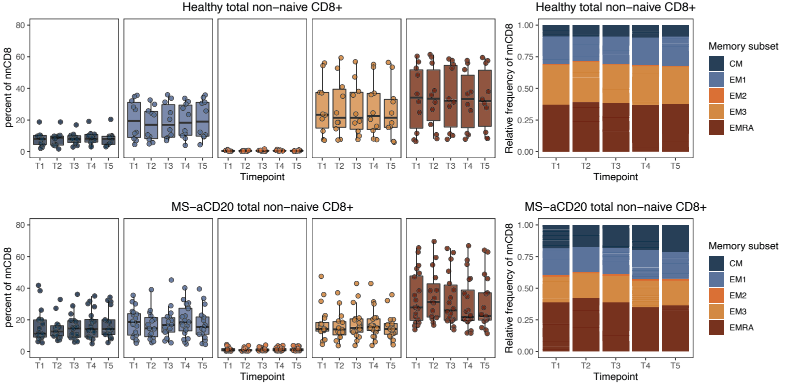

### Extended Data Figure 8.

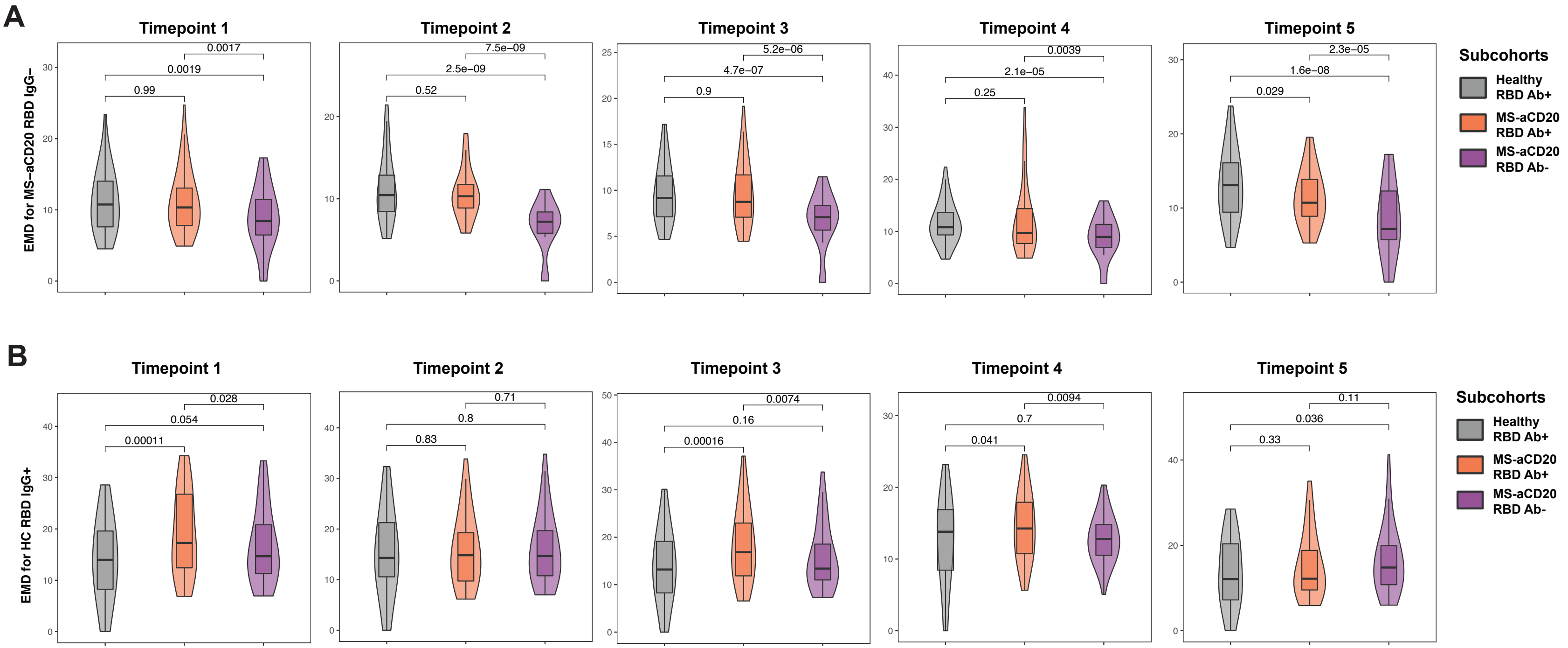
